# Intrasubject variability in Potential Early Markers of Sensorineural Hearing Damage

**DOI:** 10.1101/2024.01.18.24301474

**Authors:** Nele De Poortere, Sarineh Keshishzadeh, Hannah Keppler, Ingeborg Dhooge, Sarah Verhulst

## Abstract

The quest for noninvasive early markers for sensorineural hearing loss (SNHL) has yielded diverse measures of interest. However, comprehensive studies evaluating the test-retest reliability of multiple measures and stimuli within a single study are scarce, and a standardized clinical protocol for robust early markers of SNHL remains elusive. To address these gaps, this study explores the intra-subject variability of various potential EEG-biomarkers for cochlear synaptopathy (CS) and other SNHL-markers in the same individuals. Fifteen normal-hearing young adults underwent repeated measures of (extended high-frequency) pure-tone audiometry, speech-in-noise intelligibility, distortion-product otoacoustic emissions (DPOAEs), and auditory evoked potentials; comprising envelope following responses (EFR) and auditory brainstem responses (ABR). Results confirm high reliability in pure-tone audiometry, whereas the matrix sentence-test exhibited a significant learning effect. The reliability of DPOAEs varied across three evaluation methods, each employing distinct SNR-based criteria for DPOAE-datapoints. EFRs exhibited superior test-retest reliability compared to ABR-amplitudes. Our findings emphasize the need for careful interpretation of presumed noninvasive SNHL measures. While tonal-audiometry’s robustness was corroborated, we observed a confounding learning effect in longitudinal speech audiometry. The variability in DPOAEs highlights the importance of consistent ear probe replacement and meticulous measurement techniques, indicating that DPOAE test-retest reliability is significantly compromised under less-than-ideal conditions. As potential EEG-biomarkers of CS, EFRs are preferred over ABR-amplitudes based on the current study results.

## INTRODUCTION

In the realm of clinical application, the standard procedure for assessing auditory thresholds relies on conventional pure-tone audiometry. Consequently, studies targeting the evaluation of hearing damage related to aging, ototoxicity and excessive noise exposure have primarily centered on identifying permanent hearing threshold changes within the frequency range of 250 to 8000 Hz (Cruickshanks et al., 2010; Rabinowitz et al., 2006). Temporary threshold shifts (TTS) resulting from noise exposure were historically regarded as less concerning markers for permanent hearing damage, as indicated by the National Institute of Occupational Safety and Health (NIOSH) in 1998. However, recent rodent studies have challenged this perspective, revealing that a noise-exposure-induced TTS may coincide with permanent deficits at the synaptic level (Kujawa & Liberman, 2009b), a phenomenon referred to as cochlear synaptopathy (CS) (Fernandez et al., 2020). CS predominantly affects the connections between type-I auditory nerve fiber terminals and inner hair cells (IHCs) (Furman et al., 2013; Kujawa & Liberman, 2009a) and can result in supra-threshold temporal coding deficits (Bharadwaj et al., 2015b). Unfortunately, CS mostly affects supra-threshold sound coding without affecting routine clinical hearing sensitivity measures such as the tonal audiogram or distortion-product otoacoustic emissions (DPOAEs) (Furman et al., 2013; Lobarinas et al., 2013). Nonetheless, CS is believed to contribute to symptoms such as tinnitus and hyperacusis (Guest et al., 2017; Paul et al., 2017; Schaette & McAlpine, 2011; Wojtczak et al., 2017), and is thought to underlie other perceptual challenges, including difficulties in discriminating sounds in complex acoustic environments and impaired temporal processing of sound and speech intelligibility (Bharadwaj et al., 2015a; Garrett, 2020; Guest, Munro, Prendergast, et al., 2018; Mepani et al., 2021; Oxenham, 2016; Prendergast et al., 2017; Smith et al., 2019).

Hence, researchers persist in their quest for precise and reliable measures of early SNHL markers in clinical settings. While numerous studies have investigated the test-retest reliability of various measures, there is a noticeable lack of research that administers a comprehensive test battery to the same individuals. Moreover, as novel SNHL treatments emerge, there is a pressing need for robust biomarkers to monitor treatment effectiveness. Consequently, this study aims to explore the intra-subject variability of potential EEG biomarkers for CS and other early SNHL indicators within the same individuals, with the objective of exploring their suitability for clinical use. By conducting multiple test on the same participants, we aim to provide a unique opportunity to compare intra-subject variability.

### Early Indicators of Outer Hair Cell Loss

According to literature, extended high frequency (EHF) audiometry presents a more sensitive approach for the early detection of noise induced hearing loss (NIHL) compared to conventional frequencies (Singh et al., 2009; Wang et al., 2000). Additionally, EHF audiometry has proven to be a valuable predictive tool for identifying the risk of NIHL (Hunter et al., 2020; Mehrparvar et al., 2011), highlighting its clinical significance in the field of hearing health assessment.

An alternative, or complementary, approach to behavioral audiometry involves DPOAEs, which closely reflect the integrity of cochlear structures; particularly the outer hair cells (OHCs) (Jansen et al., 2009). Furthermore, DPOAEs are recognized for their sensitivity in detecting subtle cochlear damage before it manifests in pure-tone hearing threshold elevations (Coradini et al., 2007; Glavin et al., 2021; Knight et al., 2007; Reavis et al., 2015). Moreover, DPOAEs offer distinct advantages such as rapid acquisition, non-participatory nature, and suitability for measurement by non-specialist personnel trained in emission assessments (Reavis et al., 2015). However, despite the crucial role of DPOAEs in audiology, there remains a noticeable scarcity of studies that concurrently investigate the most suitable DPOAE evaluation methodologies, along with the reliability of auditory thresholds, DP-amplitudes, and DP-thresholds within the same cohort of subjects.

### Indicators of Cochlear Synaptopathy

Promising EEG biomarkers for CS include prominent suprathreshold neural potentials such as the Auditory Brainstem Response (ABR) wave I amplitude and the Envelope Following Response (EFR) strength, as highlighted in several studies (Guest et al., 2017; Liberman et al., 2016; Shaheen et al., 2015; Wilson et al., 2021). Animal models have emphasized the significance of ABR-amplitudes and EFR-strengths as clinical metrics for diagnosing age- and noise-induced CS (Kujawa & Liberman, 2009a; Shehabi et al., 2022; Skoe & Tufts, 2018). However, translating these measures as early biomarkers from rodents to humans faces challenges. Firstly, intersubject variabilities stemming from differences in head size and sex (Mitchell et al., 1989), electrode resistance, and various sources of electrical noise between individuals or sessions (Plack et al., 2016) act as confounding factors, limiting their diagnostic utility in humans. Secondly, while the early wave I of the ABR is thought to have diagnostic potential in individual listeners due to its clear link to CS in animal models, the OHC-loss aspect of sensorineural hearing damage also affects its amplitude (Verhulst et al., 2016). Additionally, the sensitivity of ABR measurements to low spontaneous rate (SR) ANFs, particularly vulnerable to CS, has been questioned due to the delayed onset response of these fibers (Bourien et al., 2014). ABRs are evoked by transient stimuli and primarily reflect onset responses, which tend to be relatively small in low SR fibers (Rhode & Smith, 1985; Taberner & Liberman, 2005). In contrast, EFR-strengths in response to sinusoidal amplitude modulated (SAM) tones are primarily driven by low SR-ANFs, especially when the modulation depths are shallow (Bharadwaj et al., 2014). High SR-ANFs also contribute to the EFR, when the modulation depth is maximal, as e.g. shown in the auditory simulation models of Encina-Llamas et al. (2019), Vasilkov et al. (2021), and Van Der Biest et al. (2023). Additionally, it is important to acknowledge that there are multiple sources (including AN) that can contribute to the recorded EFRs. However, the contributing sources can be targeted towards more peripheral generators (AN, CN, IC) by using higher modulation frequencies above 80 Hz, as suggested by Purcell et al. (2004).

Given that CS has been assumed to impact suprathreshold hearing sensitivity and speech recognition abilities, particularly in challenging listening conditions (Kujawa & Liberman, 2009a; Lin et al., 2011; Parthasarathy & Kujawa, 2018; Skoe et al., 2019), various speech recognition in noise tests have been employed in human studies to explore CS (Garrett, 2020; Grinn et al., 2017; Guest, Munro, & Plack, 2018; Mepani et al., 2021; Vande Maele et al., 2021). Nonetheless, research into the within-subject variability of these tests has remained limited.

In sum, the reliability of proposed noninvasive metrics as early markers of SNHL in humans remains uncertain due to challenges posed by both intra- and inter-subject variabilities, impeding integration into clinical practice. This study explores the intra-subject variability of various potential EEG biomarkers for CS and other early SNHL indicators within the same individuals across three sessions in a cohort of normal hearing listeners. The aim is to explore and compare potential early indicators of SNHL that could enhance clinical diagnostics and monitoring of SNHL.

## MATERIALS AND METHODS

### PARTICIPANTS AND STUDY DESIGN

Fifteen young adults, nine men and six women, aged between 18 and 25 years (mean age 21.0 years ± 1.77 standard deviation; SD) participated at three test sessions. Participant selection involved administering a hearing evaluation questionnaire, followed by PTA and tympanometry during the initial session. Individuals with known hearing disorders, a history of ear surgery, or tinnitus were excluded. The study encompassed three distinct sessions denoted as session 1, 2, and 3. Between each consecutive session, a time interval of two to three days was maintained, with the exception of two participants who had a 14- and 15-day interval between session 2 and 3. Throughout these intervals, participants were instructed to abstain from exposure to loud activities. During the first session, the best ear was selected based on PTA at conventional frequencies. During each session, participants completed a comprehensive test battery consisting of (EHF) PTA, speech in quiet (SPiQ)- and speech in noise (SPiN)-tests, DPOAEs and AEP-measurements. The selection for the right ear was made for 10 participants, while the left ear was tested for five participants. As part of Covid-19 safety measures, subjects wore a face mask during the measurements. The test protocol had a maximum duration of three hours, and tests were administered in a consistent sequence for all subjects across all sessions. This study received approval from the UZ Gent ethical committee (BC-05214) and adhered to the ethical principles outlined in the Declaration of Helsinki. All participants were informed about the testing procedures and provided an informed consent.

### OTOSCOPY AND TYMPANOMETRY

Otoscopy of the ear canal and the tympanic membrane was performed using a Heine Beta 200 LED otoscope (Dover, USA), and showed bilateral normal otoscopic in all subjects. Middle-ear admittance was bilaterally measured, followed by unilateral measurements (best ear) in the follow-up sessions, using a GSI TympStar (Grason-Stadler) tympanometer (Minneapolis, USA) with a 226 Hz, 85 dB sound pressure level (SPL) probe tone. All tympanograms were defined as a type-A according to the Liden-Jerger classification (Jerger, 1970; Lidén, 1969).

### PURE-TONE AUDIOMETRY

Pure-tone thresholds were determined in a double-walled sound-attenuating booth by the use of an Equinox Interacoustics audiometer (Middelfart, Denmark). Stimuli were transmitted using Interacoustics TDH-39 headphones (Middelfart, Denmark) and Sennheiser HDA-200 headphones (Wedemark, Germany) for conventional frequencies and EHFs, respectively. Air-conduction thresholds were measured using the modified Hughson-Westlake procedure at conventional octave frequencies 0.125, 0.250, 0.500, 1, 2, 4 and 8 kHz, half-octave frequencies 3 and 6 kHz, and EHF 10, 12.5, 14, 16, and 20 kHz. Both ears were tested at the first session to determine the test ear for the subsequent measurements, selecting the ear with superior thresholds on conventional frequencies. All participants were classified as having normal hearing according to the World Health Organization’s (WHO) guidelines, which define normal hearing as a better-ear audiometric threshold averaged over 0.5, 1, 2, and 4 kHz equal or below 20 dB HL (Organization, 2021).

### SPEECH INTELLIGIBILITY IN QUIET AND IN NOISE (SPiQ AND SPiN)

During each session, the SPiQ- and SPiN tests were administered in a quiet testing room, using the Flemish Matrix sentence test (Luts et al. 2014) and Apex 3 software (Francart et al., 2008). The sentences were presented to the best ear using a laptop connected to a Fireface UCX soundcard (RME) (Haimhausen, Germany) and HDA-300 (Sennheiser) headphones (Wedemark, Germany).

Speech performance was evaluated through four experimental tests lists, encompassing both broadband (BB) and high-pass filtered (HP) speech in quiet (SPiQ) and in noise (SPiN). In each session, participants were randomly assigned all four test lists, each containing 20 sentences in a randomized sequence. Due to protocol adjustments, BB-quiet was not executed for two participants. To counteract potential learning effects, two additional BB-noise training lists were administered (Luts et al., 2014).

BB-quiet involved presenting speech without filtering, while HP-quiet applied a zero-phase 1024th-order FIR HP-filter (cutoff 1650 Hz) to the speech signal. For HP-noise, both speech and noise signals were filtered using the same cutoff values as HP-quiet, whereas BB-noise had no filtering on either the speech or noise signals.

The matrix-test consisted of a corpus of 50 words, categorized into 10 names, 10 verbs, 10 numerals, 10 adjectives, and 10 nouns. All sentences shared identical syntactical structures, and the semantic content remained unpredictable. The adaptive procedure outlined by Brand & Kollmeier (2002), was employed for all test lists, implementing a staircase paradigm to ascertain the speech-reception threshold. The speech level was adjusted by a maximum of 5 dB, progressively decreasing to a minimal step size of 0.1 dB. For SPiQ, the procedure commenced at a level of 50 dB SPL, while for SPiN, the noise was maintained at a fixed level of 70 dB SPL, starting at a SNR of -4 dB. In all tests lists, subjects were instructed to repeat the five-word sentences in a forced-choice setting, with 10 options provided for each word. The mean signal level or mean SNR from the six last reversals was utilized to determine the SPiQ and SPiN thresholds.

### DISTORTION PRODUCT OTO-ACOUSTIC EMISSIONS (DPOAEs)

During each session, DPOAE measurements were carried out on the designated ear, encompassing DP-grams and DP-thresholds. DPOAEs were collected in a quiet testing room, employing the Universal Smart Box (Intelligent Hearing Systems IHS) (Miami, United States). To ensure controlled conditions, both ears were shielded using earmuffs (Busters) (Kontich, Belgium) that were placed on top of a 10D IHS OAE-probe (Miami, United States). DPOAE responses and noise amplitudes were quantified using the simultaneous presentation of two primary tones, with f1 and f2 featuring a frequency ratio f2/f1 of 1.22. Noise artifact rejection was set at 10 dB SPL, and a total of 32 sweeps were recorded for each frequency or input-output level.

DPOAE responses and noise amplitudes were obtained with a primary tone level combination of L1/L2 = 65/55 dB SPL and f2 ranging from 553 to 8837 Hz at two points per octave, and from 8837 to 11459 Hz at eight points per octave, resulting in twelve frequency bands with center frequencies 0.5, 0.7, 1.0, 1.4, 2.0, 2.8, 4.0, 5.7, 8.0, 8.7, 9.5, and 10.3 kHz.

DP-thresholds were obtained at octave frequencies between 0.5 to 8 kHz (i.e. √(f1*f2) = 501, 1000, 2000, 3998, 8001, and 10376 Hz) with L2 ranging from 35 to 70 dB SPL in steps of 5 dB. L1/L2 varied across L2 intensities using the scissor paradigm of Kummer et al. (1998) whereby L1 = 0,4 L2 + 39 dB. Extrapolation and non-linear regression were used to estimate DP-thresholds in which a cubic function was fit to the I/O functions of DPOAE measurements of each frequency following the method of Verhulst et al. (2016). This way, DP-thresholds were determined as the level of L2 at which the curve reached the distortion component of -25 dB SPL.

The evaluation of DPOAEs was subdivided into three evaluation methods using commonly used inclusion criteria, i.e. response amplitude ≥ the noise floor; response amplitude ≥ 2SD above the noise floor; response amplitude ≥ 6 dB above the noise floor. When responses did not meet the inclusion criteria, the amplitudes were set to the noise floor levels. DP-thresholds outside the range -10 – 60 dB (Boege & Janssen, 2002) were excluded, since these responses are not considered as valid.

### AUDITORY EVOKED POTENTIAL (AEP) MEASUREMENTS

AEP measurements, including EFRs and ABRs were conducted at the test ear using the IHS universal Smart box and SEPCAM software (Miami, United States). Recordings were performed in a quiet testing room, with subjects seated in a reclining chair, watching a muted video while resting their heads on a soft pillow. To minimize alpha-wave interference, subjects were instructed to relax without falling asleep. Controlled conditions within the hospital setting were maintained by shielding both ears with earmuffs (Busters) (Kontich, Belgium), turning off extraneous lights and electronic devices, and applying NuPrep gel for skin preparation. Disposable Ambu Neuroline electrodes (Ballerup, Denmark) were placed on the vertex (inverting electrode), nasal flank on the non-test ear side (ground electrode), and bilateral mastoids (non-inverting electrodes). Electrode impedances were kept below 3 kΩ, and auditory stimuli were presented using etymotic ER-2 ear-probes (Chicago, USA).

EFRs were evoked using two stimulus types, distinguished by their modulation waveform, i.e. a sinusoidal amplitude modulated (SAM)-stimulus with a carrier frequency of 4 kHz, and rectangularly amplitude modulated (RAM)-stimuli, with carrier frequencies 4 and 6 kHz, and a duty cycle of 25% (Van Der Biest et al., 2023; Vasilkov et al., 2021). EFRs were evoked using 1000 alternating polarity sweeps. Stimuli had a modulation frequency of 110 Hz, a modulation depth of 100% and a duration of 500 ms which were presented at a rate of 2 Hz. The RAM stimuli with different carriers were calibrated in such a way to have the same peak-to-peak amplitude as a 70 dB SPL SAM-tone (carrier: 4 kHz, modulation frequency: 110 Hz, modulation depth: 100%). In this regard, the calibrated RAM stimuli with different carrier frequencies were presented at 68.24 dB SPL and had the same peak-to-peak amplitudes.

The EFR processing was performed in Matlab. Firstly, the recordings were filtered using a bandpass filter with low and high cutoff frequencies of 30 Hz and 1500 Hz, respectively. After filtering the EFRs, epoching and baseline correction was performed. Lastly, a bootstrapping approach according to Zhu et al. (2013) was adopted in the frequency domain to estimate the noise-floor and variability of the EFR, as detailed in Keshishzadeh et al. (2020). Subsequently, EFR-strengths represented the summation of the signal-to-noise spectral magnitude at the fundamental frequency and its following three harmonics, i.e. 110, 220, 330 and 440 Hz (Vasilkov et al., 2021).

ABRs were evoked using 4000 alternating polarity sweeps of six stimulus types, i.e. three broadband 80-µs clicks presented at levels of 70, 80 and 90 dBpeSPL and three narrowband toneburst (TB)-stimuli at 0.5 kHz, 1 kHz and 4 kHz with a stimulus duration of 5 ms, 4 ms and 2 ms, respectively. Clicks were presented at a rate of 11 Hz and TBs had a rate of 20 Hz. ABR recordings were filtered offline between 100 and 1500 Hz using a zero-phase filter. Afterwards, epoching and baseline correction was performed akin to the method described for EFR processing. After baseline correction, epochs were averaged to yield the ABR waveform. ABR waves I, III and V were manually peak-picked by audiologists to identify the respective ABR amplitudes (µV) and latencies (ms). ABR amplitudes were defined peak to baseline.

### STATISTICAL ANALYSIS

All statistical analyses were conducted using IBM SPSS Statistics 27. The data-analysis employed a four-tiered methodology, including one-way repeated measures ANOVA, two-way random average measures intraclass correlation coefficient (ICC), standard errors of measurement (SEM), and calculation of individual 95% confidence intervals (95%CI). Firstly, a One-way repeated measures ANOVA was employed to assess variations in PTA, SPiQ and SPiN, DPOAEs and AEP outcomes across three consecutive measurements. Descriptive statistics were calculated, and tests for normality, including the Shapiro-Wilk test, histograms, Q-Q plots, and box-and-whisker plots, were conducted to evaluate the assumptions for the one-way repeated measures ANOVA. When a significance level of p < 0.05 was reached, post-hoc analysis using least-square means was performed to assess intersession differences. A two-tailed significance level of p < 0.017 was used, adjusted by Bonferroni correction for multiple comparisons. This correction effectively controls for multiple comparisons within individual parameters tested across three sessions; however, it does not fully address the risk of Type I errors across the extensive range of parameters analyzed. This methodological choice was made to balance the exploratory nature of the study with the need for statistical rigor, focusing primarily on minimizing Type II errors to ensure that genuine effects were not overlooked. Secondly, two-way random-average-measures intraclass correlation coefficients were computed to determine the relative consistency, i.e. the consistency of the position of individual scores relative to others. The interpretation of ICC-values followed the classification system proposed by Koo and Li (2016): excellent ICC (>0.90), good ICC (0.75 - 0.90), moderate ICC (0.50 – 0.75) and poor ICC (< 0.50). Thirdly, SEM-scores were calculated to represent the reliability within repeated measures for an individual subject, reflecting absolute consistency. The latter is calculated as SEM = s*√(1-ICC), where ‘s’ represents the standard deviation of all measurements. Finally, given that the substantial intersubject variability observed in each measure had an influence on the group-based test-retest 95% confidence intervals (CIs), we additionally computed 95%CIs of the repeated measures for each individual separately to visually assess the reliability of different hearing parameters in comparison to each other. This process entailed calculating 95% CIs across measurement sessions for each parameter and subject. The resulting distribution of individual CIs across subjects, is visualized using Kernel Density Estimation (KDE) plots, representing the upper and lower bounds of obtained test-retest CIs for each measure and each subject. These KDE-plots served to illustrate the variability of test-retest CIs across subjects and enhance the interpretation of test-retest variations within the data.

## RESULTS

### PURE-TONE AUDIOMETRY

At session 1, the mean pure-tone average at 3, 4, 6 and 8 kHz (PTA3-8kHz) was 8.08 dB HL (SD 6.663, range -2.00 – 18.00) and the mean pure-tone average at 10, 12.5, 14, 16 and 20 kHz (PTA EHF10-20 kHz) was 4.40 dB HL (SD 8.382, range -8.00 – 18.00). One-way repeated measures ANOVA revealed no significant changes in pure-tone thresholds between measurements, except for the 0.25 kHz auditory thresholds [F(2, 28) = 6.526, *p* = .005]. Pairwise comparisons indicated a significant change of -4.00 dB from session 1 to session 3 (*p* = .009). Table I presents the averages per session and frequency, along with ICCs and SEMs for each tested frequency. In general, good to excellent ICCs with highly significant between subjects reliability (*p* < .001) were obtained and small SEMs were observed. However, at 6 kHz, 8 kHz and 20 kHz, lower ICCs were observed alongside wider corresponding 95%CIs and higher SEMs.

**Table I.**
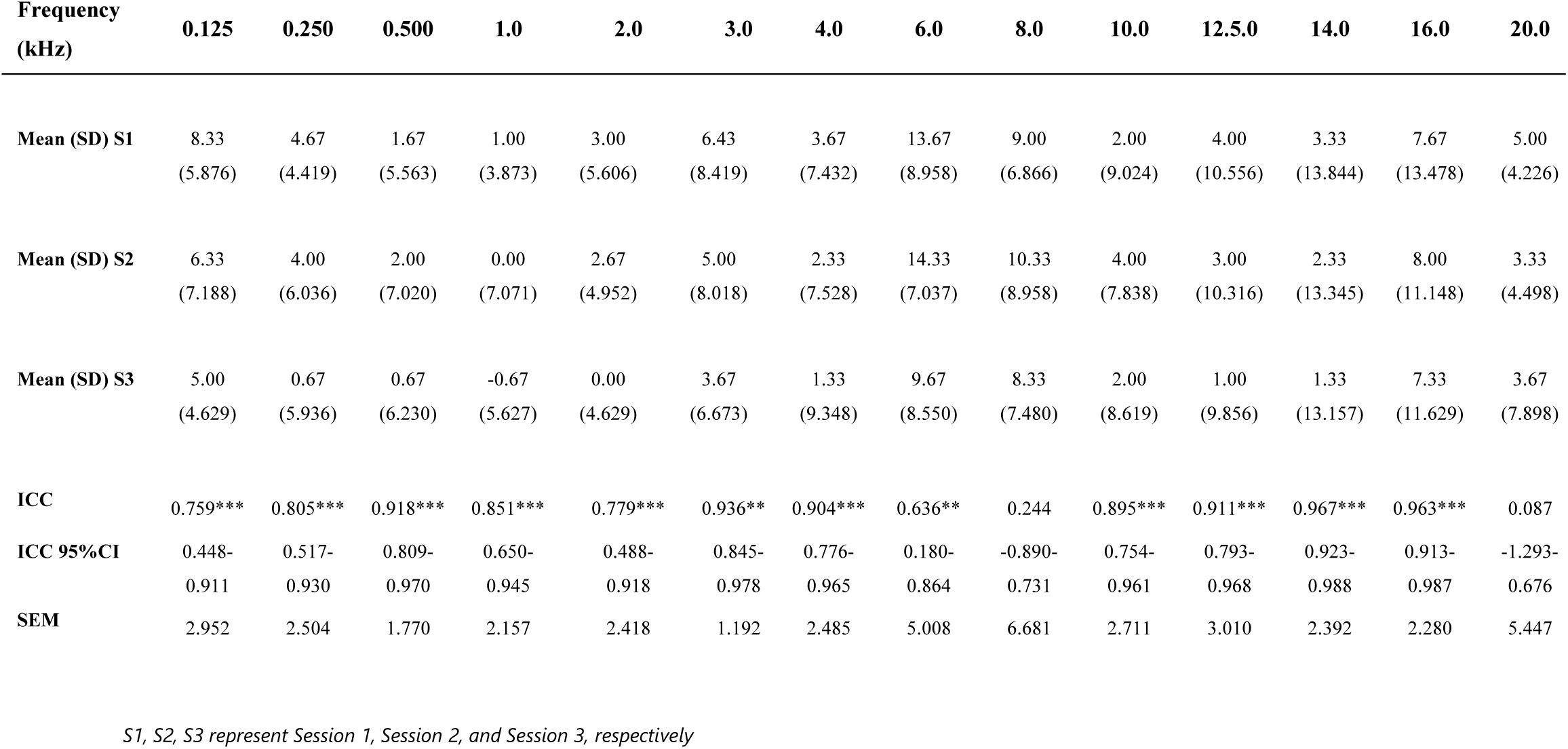
Summary of audiometric threshold averages (dB HL) per session and frequency, alongside Intraclass Correlation Coefficients (ICCs) and corresponding 95% Confidence Intervals (CIs), and Standard Error of Measurements (SEMs) for all tested frequencies. P-values for between-subjects variability are reflected as *(0.05 > p > 0.01), **(0.01 > p > 0.001), and ***(p < 0.001).

| Frequency<br>(kHz) | 0.125 | 0.250 | 0.500 | 1.0 | 2.0 | 3.0 | 4.0 | 6.0 | 8.0 | 10.0 | 12.5.0 | 14.0 | 16.0 | 20.0 |
| --- | --- | --- | --- | --- | --- | --- | --- | --- | --- | --- | --- | --- | --- | --- |
| **Mean (SD) S1** | 8.33<br>(5.876) | 4.67<br>(4.419) | 1.67<br>(5.563) | 1.00<br>(3.873) | 3.00<br>(5.606) | 6.43<br>(8.419) | 3.67<br>(7.432) | 13.67<br>(8.958) | 9.00<br>(6.866) | 2.00<br>(9.024) | 4.00<br>(10.556) | 3.33<br>(13.844) | 7.67<br>(13.478) | 5.00<br>(4.226) |
| **Mean (SD) S2** | 6.33<br>(7.188) | 4.00<br>(6.036) | 2.00<br>(7.020) | 0.00<br>(7.071) | 2.67<br>(4.952) | 5.00<br>(8.018) | 2.33<br>(7.528) | 14.33<br>(7.037) | 10.33<br>(8.958) | 4.00<br>(7.838) | 3.00<br>(10.316) | 2.33<br>(13.345) | 8.00<br>(11.148) | 3.33<br>(4.498) |
| **Mean (SD) S3** | 5.00<br>(4.629) | 0.67<br>(5.936) | 0.67<br>(6.230) | -0.67<br>(5.627) | 0.00<br>(4.629) | 3.67<br>(6.673) | 1.33<br>(9.348) | 9.67<br>(8.550) | 8.33<br>(7.480) | 2.00<br>(8.619) | 1.00<br>(9.856) | 1.33<br>(13.157) | 7.33<br>(11.629) | 3.67<br>(7.898) |
| **ICC** | 0.759*** | 0.805*** | 0.918*** | 0.851*** | 0.779*** | 0.936** | 0.904*** | 0.636** | 0.244 | 0.895*** | 0.911*** | 0.967*** | 0.963*** | 0.087 |
| **ICC 95%CI** | 0.448-0.911 | 0.517-0.930 | 0.809-0.970 | 0.650-0.945 | 0.488-0.918 | 0.845-0.978 | 0.776-0.965 | 0.180-0.864 | -0.890-0.731 | 0.754-0.961 | 0.793-0.968 | 0.923-0.988 | 0.913-0.987 | -1.293-0.676 |
| **SEM** | 2.952 | 2.504 | 1.770 | 2.157 | 2.418 | 1.192 | 2.485 | 5.008 | 6.681 | 2.711 | 3.010 | 2.392 | 2.280 | 5.447 |

*S1, S2, S3 represent Session 1, Session 2, and Session 3, respectively*

### SPEECH IN QUIET AND SPEECH IN NOISE (SPiQ AND SPiN)

The distribution of SPiQ (dB SPL) and SPiN (dB SNR) thresholds across subjects is depicted in Figures 1A and 1B, respectively.

**Figure 1:**
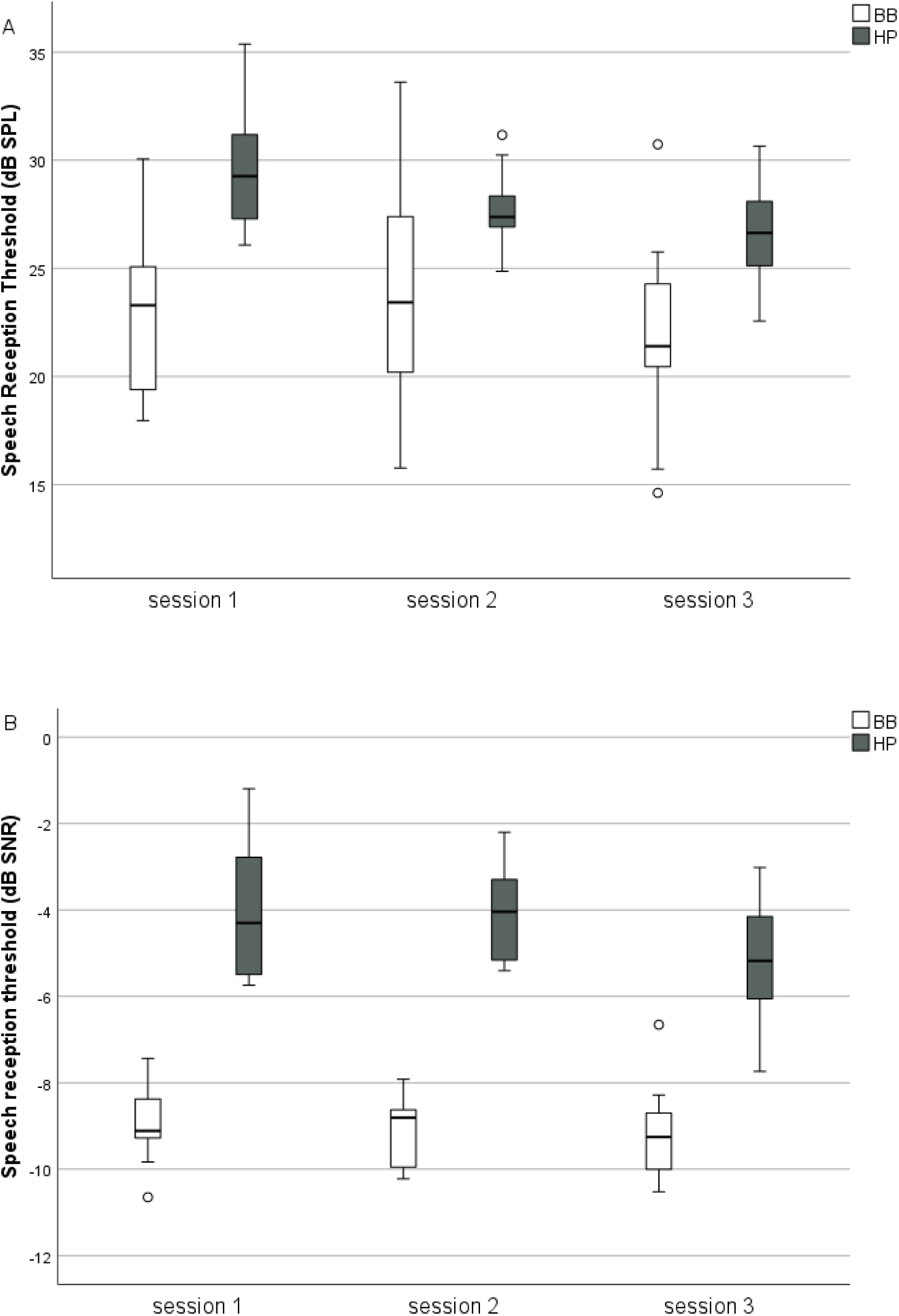
Boxplots illustrating the distribution of SPiQ (A) and SPiN (B) thresholds across sessions. White boxplots represent speech audiometry thresholds for BB-stimuli, while grey boxplots indicate thresholds for HP-stimuli.

One-way repeated measures ANOVA indicated no significant alterations in thresholds between measurement sessions for BB-quiet [F(2, 24) = 1.549, *p* > 0.05], nor BB-noise [F(2, 28) = 0.690, *p* > 0.05]. In contrast, significant threshold changes were found for HP-quiet [F(2, 28) = 12.266, *p* < 0.001], and HP-noise [F(2, 28) = 7.788, *p* = 0.002]. Pairwise comparisons unveiled threshold improvements for HP-quiet between each session with non-significant changes of 1.72 dB SPL from session 1 to session 2 (*p* > 0.017) and 1.12 dB SPL from session 2 to session 3 (*p* > 0.017), and a significant change of 2.84 dB SPL from session 1 to session 3 (*p* = 0.004). Furthermore, a significant change of 1.13 dB SNR between session 2 and 3 (*p* = 0.002), and a significant change of 1.21 dB SNR between session 1 and 3 (*p* = 0.010) were found for HP-noise. It should be noted that the initial training procedure before the start of the measurements only incorporated BB-speech in noise. Table II displays ICCs and SEMs for SPiQ and SPiN thresholds, indicating moderate ICCs with highly significant between-subject variability.

**Table II.** Summary of broadband (BB) and high-pass filtered (HP) speech audiometry threshold in quiet (dB SPL) and in noise (dB SNR) averages per session, along with Intraclass Correlation Coefficients (ICCs) and corresponding 95% Confidence Intervals (CIs), and Standard Error of Measurements (SEMs). P-values for between-subjects variability are reflected as *(0.05 > p > 0.01), **(0.01 > p > 0.001), and ***(p < 0.001).

|  | Speech audiometry in quiet |  | Speech audiometry in noise |  |
| --- | --- | --- | --- | --- |
|  | BB | HP | BB | HP |
| **Mean (SD) S1** | 22.93 (4.011) | 29.48 (2.906) | -8.93 (0.806) | -4.01 (1.522) |
| **Mean (SD) S2** | 23.69 (4.794) | 27.76 (1.763) | -9.15 (0.783) | -4.09 (1.091) |
| **Mean (SD) S3** | 21.93 (4.036) | 26.64 (2.166) | -9.22 (1.012) | -5.22 (1.424) |
| **ICC** | 0.686** | 0.666*** | 0.619** | 0.694*** |
| **ICC 95%CI** | 0.204-0.900 | 0.196-0.878 | 0.096-0.862 | 0.286-0.887 |
| **SEM** | 2.395 | 1.481 | 0.532 | 0.797 |

*S1, S2, S3 represent Session 1, Session 2, and Session 3, respectively*

### DISTORTION PRODUCT OTOACOUSTIC EMISSIONS (DPOAEs)

#### DP-GRAMS

Per criterium, i.e. SNR ≥ 0, SNR ≥ 2 SD, and SNR ≥ 6 dB, one-way repeated measures ANOVA indicated no significant changes in DPOAE response amplitudes (*p* > 0.05) for all tested frequencies. The corresponding ICCs and SEMs are presented in Table III, along with the respective DPOAE noise amplitudes, which are reported only once as they remain consistent across all criteria. Overall, the SNR ≥ 0-criterium showed the highest ICCs (moderate-to-good), followed by the SNR ≥ 6 dB-criterium and the SNR ≥ 2SD-criterium, respectively. The latter criterium is additionally characterized by greater variability among the different tested frequencies. Secondly, remarkably worse ICCs were found for the lower frequencies of 1 and 1.5 kHz. SEMs showed relatively large values overall, with the SNR ≥ 2SD-criterium showing the largest values relative to the other criteria. Figures 2 A, B, and C depict KDE-plots of the zero-criterion, illustrating the distribution of individual test-retest 95% CIs for different measures. A sharp peak in the KDE signifies a more concentrated distribution of test-retest CIs, indicating good overall reliability across the test population. Conversely, a broader peak implies increased variability of individual test-retest CIs across individuals, reflecting a lower reliability for the corresponding parameter across the population.

**Figure 2:**
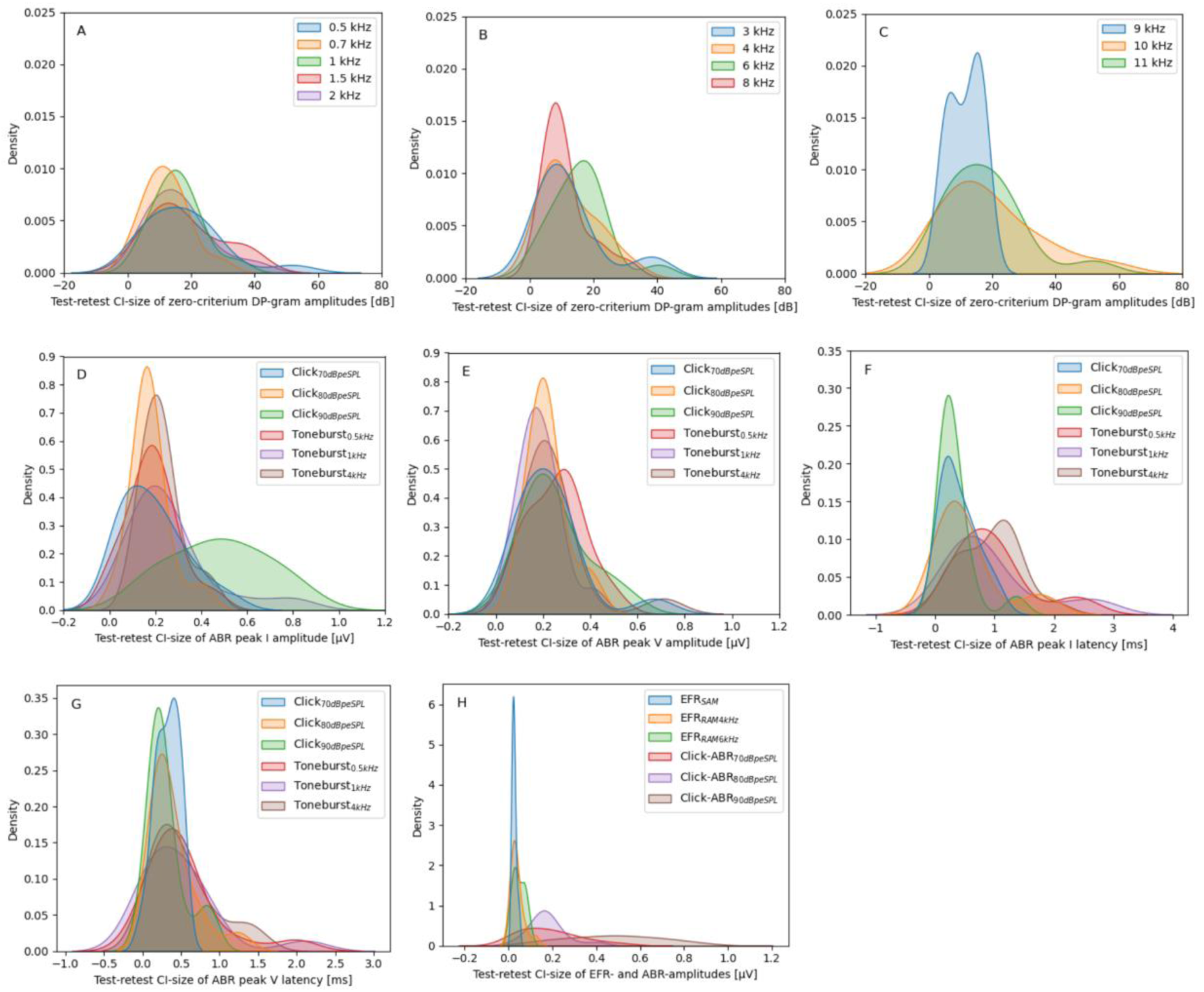
(color online) Kernel Density Estimate plots showing the distribution of individual test-retest 95% Confidence Intervals (CIs). Panels A, B and C illustrate individual test-retest 95%CIs for zero-criterion DP-grams across different frequency ranges; 0.5-2 kHz (A), 3-8 kHz (B), 9-11 kHz (C). Subsequently, Panels D-G present ABR amplitudes for wave I (D) and wave V (E), along with ABR latencies for wave I (F) and wave V (G). Additionally, panel (H) contrasts the distribution of individual 95%CIs for EFR strengths with click-ABR amplitudes. A sharp peak observed in the KDE indicates a more concentrated distribution of test-retest CIs, suggesting good overall reliability across the tested population. Conversely, a broader peak implies signifies increased variability of individual test-retest CIs across individuals, reflecting a lower reliability for the corresponding parameter across the test population.

**Table III.**
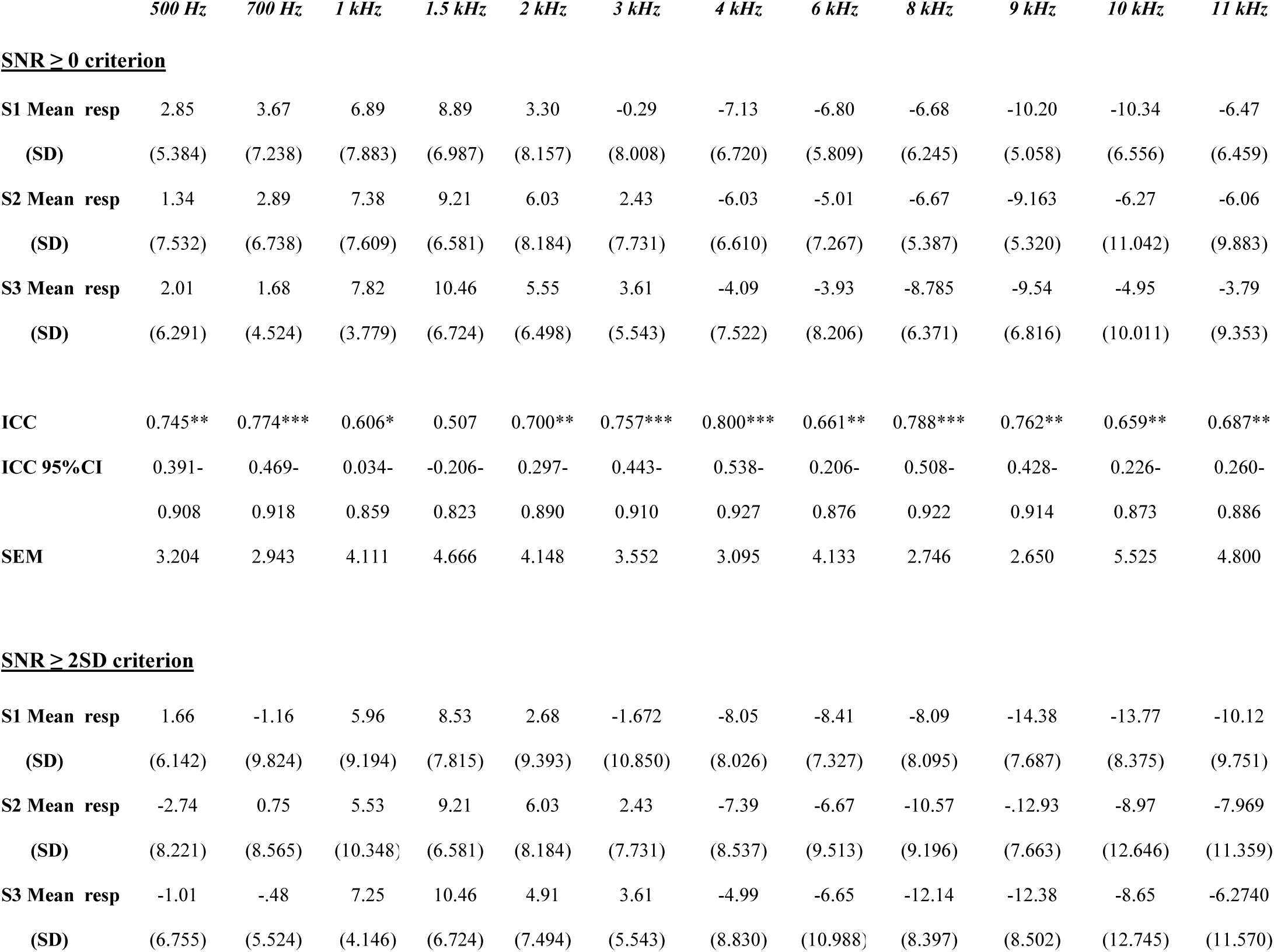
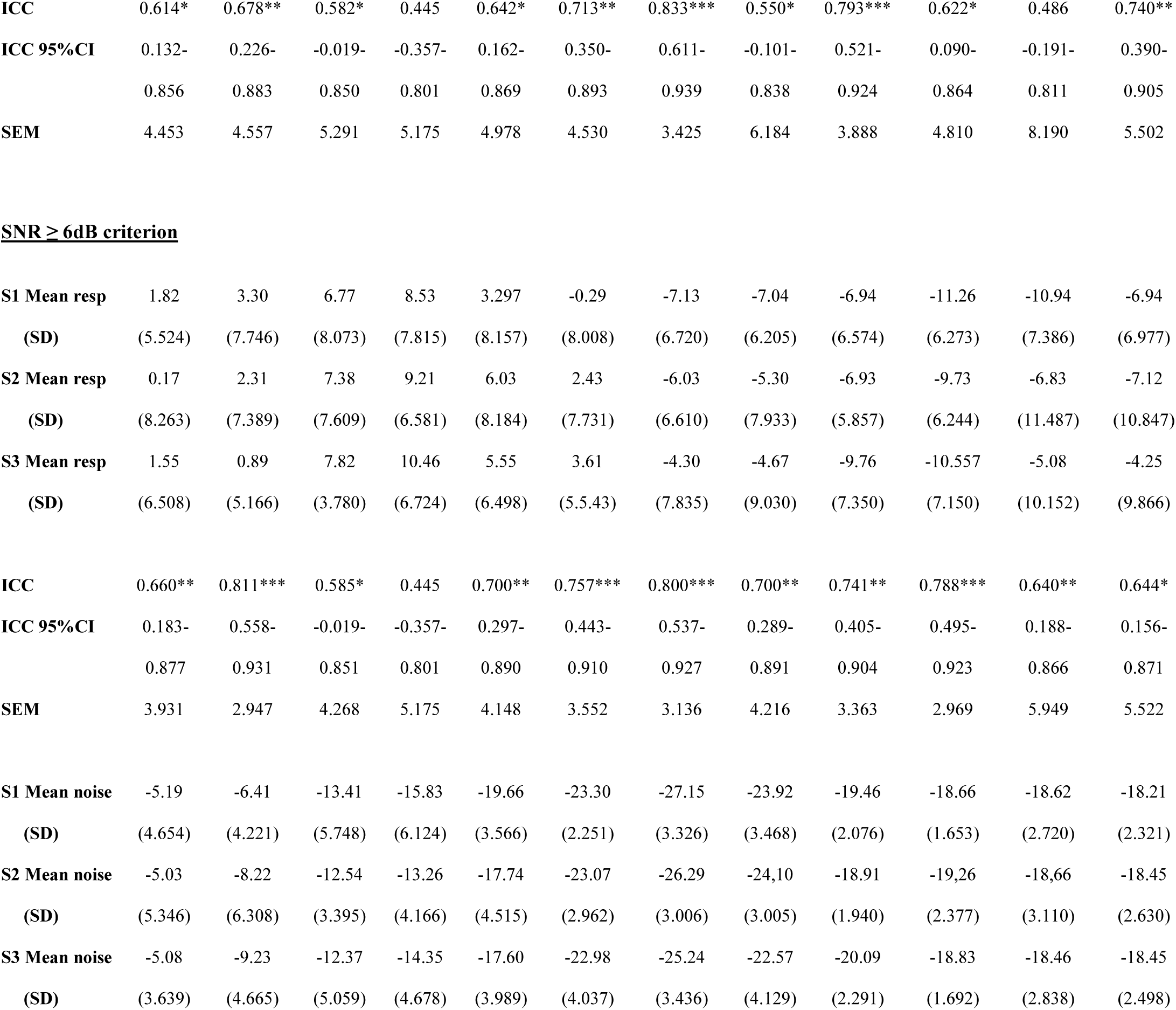
Summary of DPOAE responses (resp) and noise amplitudes (dB SPL) per session and frequency, alongside Intraclass Correlation Coefficients (ICCs) and corresponding 95% Confidence Intervals (CIs), Standard Error of Measurements (SEMs) and noise floors (dB SPL). P-values for between-subjects variability are reflected as *(0.05 > p > 0.01), **(0.01 > p > 0.001), and ***(p < 0.001).

#### DP-THRESHOLDS

One-way repeated measures ANOVA indicated no significant changes (p > 0.05) across all tested frequencies, assessed per inclusion criterion. ICCs and SEMs are shown in Table IV. The SNR ≥ 6dB-criterium showed overall the highest ICCs, followed by SNR ≥ 2SD and SNR ≥ 0, retaining both very similar ICCs and SEMs. However, overall very poor ICCs with notably wide corresponding 95%CIs, alongside large SEMs, were observed across all six tested frequencies for DPOAE thresholds.

**Table IV.**
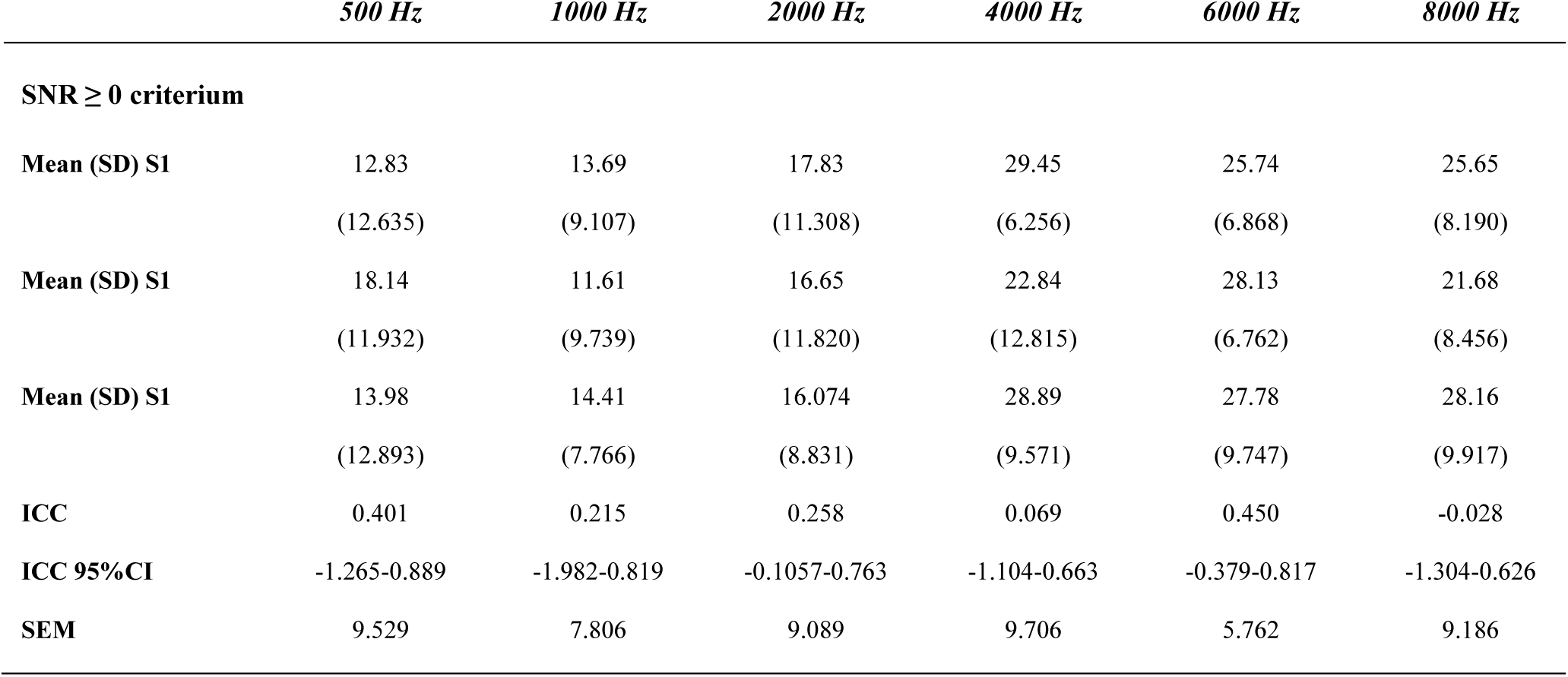
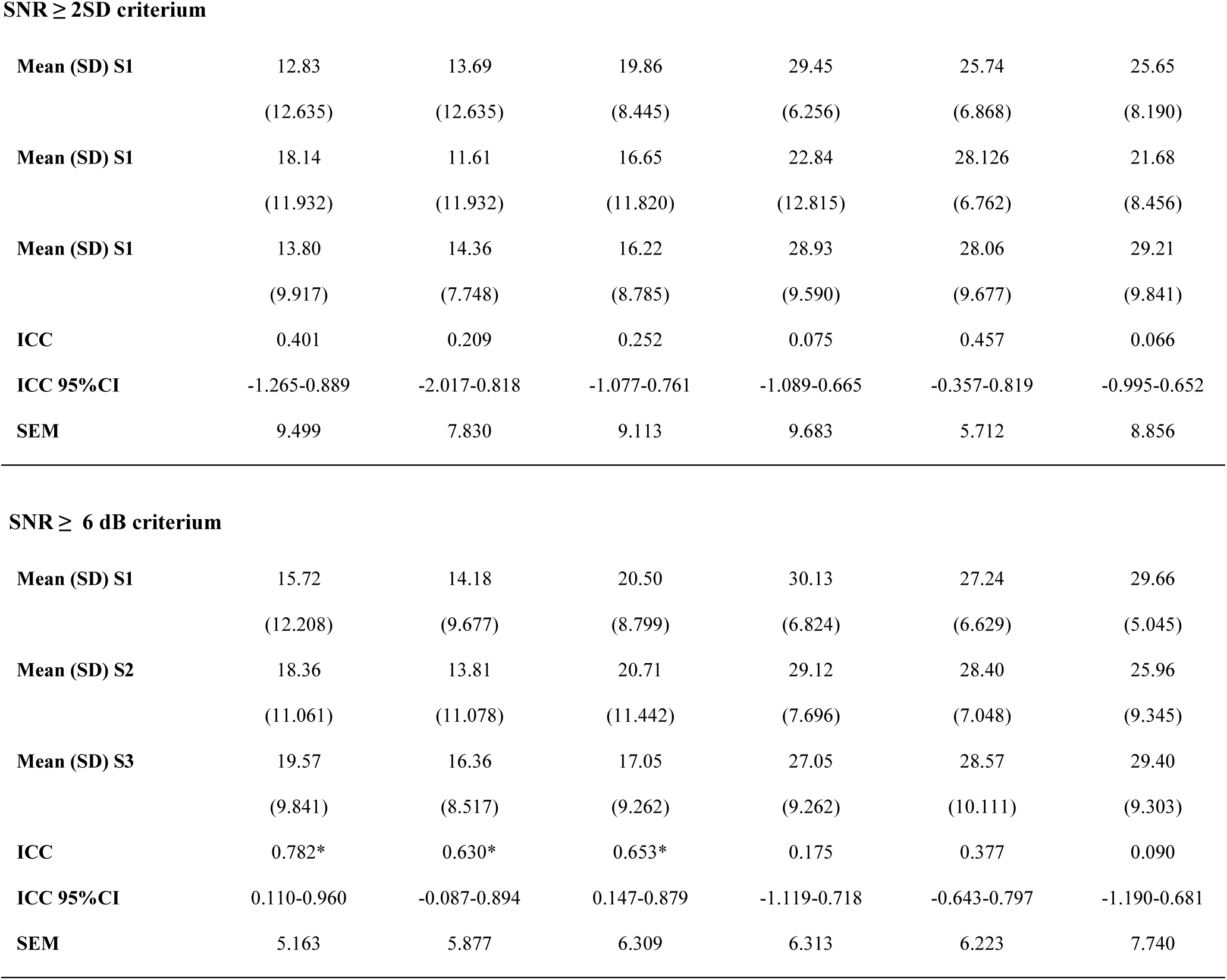
Summary of DP-thresholds amplitude averages (dB SPL) per session and frequency, alongside Intraclass Correlation Coefficients (ICCs) and corresponding 95% Confidence Intervals (CIs), and Standard Error of Measurements (SEMs). P-values for between-subjects variability are reflected as *(0.05 > p > 0.01), **(0.01 > p > 0.001), and ***(p < 0.001).

|  | *500 Hz* | *1000 Hz* | *2000 Hz* | *4000 Hz* | *6000 Hz* | *8000 Hz* |
| --- | --- | --- | --- | --- | --- | --- |
| **SNR $\geq 0$ criterium** |  |  |  |  |  |  |
| **Mean (SD) S1** | 12.83<br>(12.635) | 13.69<br>(9.107) | 17.83<br>(11.308) | 29.45<br>(6.256) | 25.74<br>(6.868) | 25.65<br>(8.190) |
| **Mean (SD) S1** | 18.14<br>(11.932) | 11.61<br>(9.739) | 16.65<br>(11.820) | 22.84<br>(12.815) | 28.13<br>(6.762) | 21.68<br>(8.456) |
| **Mean (SD) S1** | 13.98<br>(12.893) | 14.41<br>(7.766) | 16.074<br>(8.831) | 28.89<br>(9.571) | 27.78<br>(9.747) | 28.16<br>(9.917) |
| **ICC** | 0.401 | 0.215 | 0.258 | 0.069 | 0.450 | -0.028 |
| **ICC 95%CI** | -1.265-0.889 | -1.982-0.819 | -0.1057-0.763 | -1.104-0.663 | -0.379-0.817 | -1.304-0.626 |
| **SEM** | 9.529 | 7.806 | 9.089 | 9.706 | 5.762 | 9.186 |
| **SNR ≥ 2SD criterium** |  |  |  |  |  |  |
| **Mean (SD) S1** | 12.83<br>(12.635) | 13.69<br>(12.635) | 19.86<br>(8.445) | 29.45<br>(6.256) | 25.74<br>(6.868) | 25.65<br>(8.190) |
| **Mean (SD) S1** | 18.14<br>(11.932) | 11.61<br>(11.932) | 16.65<br>(11.820) | 22.84<br>(12.815) | 28.126<br>(6.762) | 21.68<br>(8.456) |
| **Mean (SD) S1** | 13.80<br>(9.917) | 14.36<br>(7.748) | 16.22<br>(8.785) | 28.93<br>(9.590) | 28.06<br>(9.677) | 29.21<br>(9.841) |
| **ICC** | 0.401 | 0.209 | 0.252 | 0.075 | 0.457 | 0.066 |
| **ICC 95%CI** | -1.265-0.889 | -2.017-0.818 | -1.077-0.761 | -1.089-0.665 | -0.357-0.819 | -0.995-0.652 |
| **SEM** | 9.499 | 7.830 | 9.113 | 9.683 | 5.712 | 8.856 |
| **SNR ≥ 6 dB criterium** |  |  |  |  |  |  |
| **Mean (SD) S1** | 15.72<br>(12.208) | 14.18<br>(9.677) | 20.50<br>(8.799) | 30.13<br>(6.824) | 27.24<br>(6.629) | 29.66<br>(5.045) |
| **Mean (SD) S2** | 18.36<br>(11.061) | 13.81<br>(11.078) | 20.71<br>(11.442) | 29.12<br>(7.696) | 28.40<br>(7.048) | 25.96<br>(9.345) |
| **Mean (SD) S3** | 19.57<br>(9.841) | 16.36<br>(8.517) | 17.05<br>(9.262) | 27.05<br>(9.262) | 28.57<br>(10.111) | 29.40<br>(9.303) |
| **ICC** | 0.782* | 0.630* | 0.653* | 0.175 | 0.377 | 0.090 |
| **ICC 95%CI** | 0.110-0.960 | -0.087-0.894 | 0.147-0.879 | -1.119-0.718 | -0.643-0.797 | -1.190-0.681 |
| **SEM** | 5.163 | 5.877 | 6.309 | 6.313 | 6.223 | 7.740 |

### DP-GRAMS AND DP-THRESHOLDS IN RELATION TO PURE-TONE AUDITORY THRESHOLDS

ICCs and SEMs of DP-grams and DP-thresholds (dB SPL), in relation to pure tone thresholds (dB HL) are illustrated in Figures 3A, B, C, and D.

**Figure 3A, 3B, 3C, and 3D:**
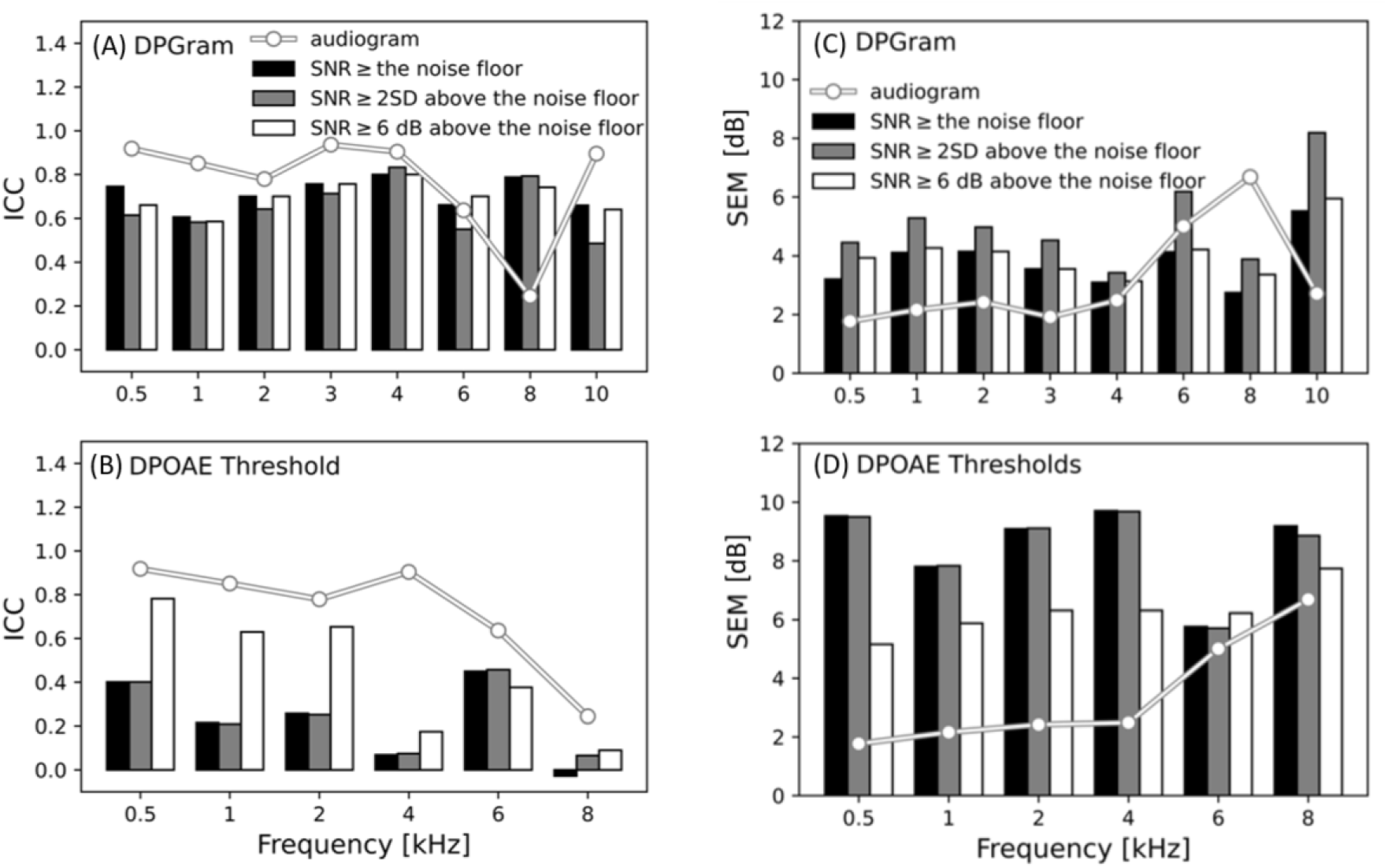
Intraclass Correlation Coefficients (ICCs) (A and B) and Standard Error of Measurements (SEMs) (C and D) of DP-amplitudes and DP-thresholds in relation to audiogram thresholds, are illustrated in panels A, B, C and D, respectively. ICCs and SEMs of the three different inclusion criteria regarding signal to noise ratio, i.e. SNR ≥ the noise floor, SNR ≥ 2SD above the noise floor, SNR ≥ 6 dB above the noise floor, are illustrated in black, grey and white, while ICCs and SEMs of pure-tone audiometry are illustrated by a line.

In terms of DP-amplitude ICCs (A), a pattern of generally lower but more consistent outcomes across different frequencies and evaluation criteria was observed compared to audiogram thresholds. Notably, exceptions were observed at 6 kHz and, predominantly, 8 kHz, where DP-amplitudes exhibited better ICCs than audiogram thresholds. This trend corresponded with the DP-amplitude SEMs (C). The DP-thresholds exhibited notably lower ICCs when juxtaposed with audiogram thresholds (B). Moreover, increased variability across different evaluation criteria was evident, particularly with improved outcomes for the 6 dB criterion. SEMs confirmed less favorable results for DP-thresholds (D), emphasizing the 6 dB criterion as the most reliable in this study population.

### AUDITORY EVOKED POTENTIALS (AEP)

#### AUDITORY BRAINSTEM RESPONSES (ABR)

A one-way repeated measures ANOVA revealed no significant differences in click-ABR amplitudes and latencies of waves I, III, and V at 70 dBpeSPL, 80 dBpeSPL, and 90 dBpeSPL, as well as for TB-stimuli, across measurement sessions (p > 0.05). Tables V and VI display Click- and TB-ABR amplitudes and latencies per session and stimulus level, alongside ICCs and SEMs. It is important to highlight that a correction of 1 ms should be applied when comparing the current latencies with those obtained using clinical-grade equipment. Generally, good-to-excellent ICCs with highly significant between-subject variances and small SEMs were observed for click-ABR wave I-, III- and V-latencies. These findings align with click-ABR wave V-amplitudes, showing good ICCs with highly significant between-subject variances, except for the click-ABR 70 dBpeSPL, retaining a moderate ICC. In contrast to wave I- and III-click latencies, wave I- and III-amplitudes showed moderate-to-very poor ICCs, with poor average measures.

**Table V.**
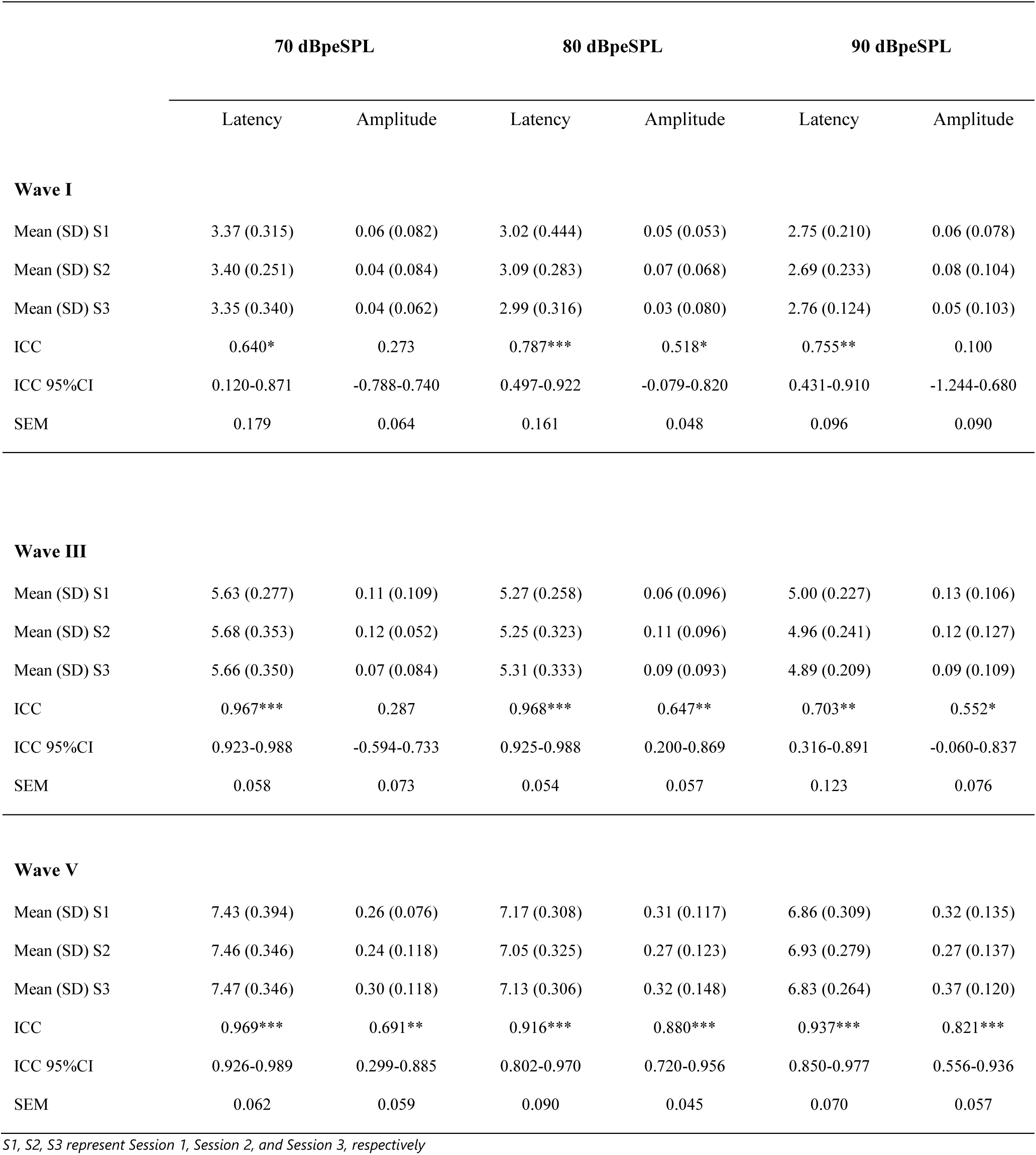
Summary of Click-ABR amplitudes (μV) and latencies (ms) per session and stimulus level (dBpeSPL), alongside Intraclass Correlation Coefficients (ICCs) and corresponding 95% Confidence Intervals (CIs), and Standard Errors of Measurements (SEMs). P-values for between-subjects variability are reflected as *(0.05 > p > 0.01), **(0.01 > p > 0.001), and ***(p < 0.001).

|  | 70 dBpeSPL |  | 80 dBpeSPL |  | 90 dBpeSPL |  |
| --- | --- | --- | --- | --- | --- | --- |
|  | Latency | Amplitude | Latency | Amplitude | Latency | Amplitude |
| **Wave I** |  |  |  |  |  |  |
| Mean (SD) S1 | 3.37 (0.315) | 0.06 (0.082) | 3.02 (0.444) | 0.05 (0.053) | 2.75 (0.210) | 0.06 (0.078) |
| Mean (SD) S2 | 3.40 (0.251) | 0.04 (0.084) | 3.09 (0.283) | 0.07 (0.068) | 2.69 (0.233) | 0.08 (0.104) |
| Mean (SD) S3 | 3.35 (0.340) | 0.04 (0.062) | 2.99 (0.316) | 0.03 (0.080) | 2.76 (0.124) | 0.05 (0.103) |
| ICC | 0.640* | 0.273 | 0.787*** | 0.518* | 0.755** | 0.100 |
| ICC 95%CI | 0.120-0.871 | -0.788-0.740 | 0.497-0.922 | -0.079-0.820 | 0.431-0.910 | -1.244-0.680 |
| SEM | 0.179 | 0.064 | 0.161 | 0.048 | 0.096 | 0.090 |
| **Wave III** |  |  |  |  |  |  |
| Mean (SD) S1 | 5.63 (0.277) | 0.11 (0.109) | 5.27 (0.258) | 0.06 (0.096) | 5.00 (0.227) | 0.13 (0.106) |
| Mean (SD) S2 | 5.68 (0.353) | 0.12 (0.052) | 5.25 (0.323) | 0.11 (0.096) | 4.96 (0.241) | 0.12 (0.127) |
| Mean (SD) S3 | 5.66 (0.350) | 0.07 (0.084) | 5.31 (0.333) | 0.09 (0.093) | 4.89 (0.209) | 0.09 (0.109) |
| ICC | 0.967*** | 0.287 | 0.968*** | 0.647** | 0.703** | 0.552* |
| ICC 95%CI | 0.923-0.988 | -0.594-0.733 | 0.925-0.988 | 0.200-0.869 | 0.316-0.891 | -0.060-0.837 |
| SEM | 0.058 | 0.073 | 0.054 | 0.057 | 0.123 | 0.076 |
| **Wave V** |  |  |  |  |  |  |
| Mean (SD) S1 | 7.43 (0.394) | 0.26 (0.076) | 7.17 (0.308) | 0.31 (0.117) | 6.86 (0.309) | 0.32 (0.135) |
| Mean (SD) S2 | 7.46 (0.346) | 0.24 (0.118) | 7.05 (0.325) | 0.27 (0.123) | 6.93 (0.279) | 0.27 (0.137) |
| Mean (SD) S3 | 7.47 (0.346) | 0.30 (0.118) | 7.13 (0.306) | 0.32 (0.148) | 6.83 (0.264) | 0.37 (0.120) |
| ICC | 0.969*** | 0.691** | 0.916*** | 0.880*** | 0.937*** | 0.821*** |
| ICC 95%CI | 0.926-0.989 | 0.299-0.885 | 0.802-0.970 | 0.720-0.956 | 0.850-0.977 | 0.556-0.936 |
| SEM | 0.062 | 0.059 | 0.090 | 0.045 | 0.070 | 0.057 |

*S1, S2, S3 represent Session 1, Session 2, and Session 3, respectively*

**Table VI.**
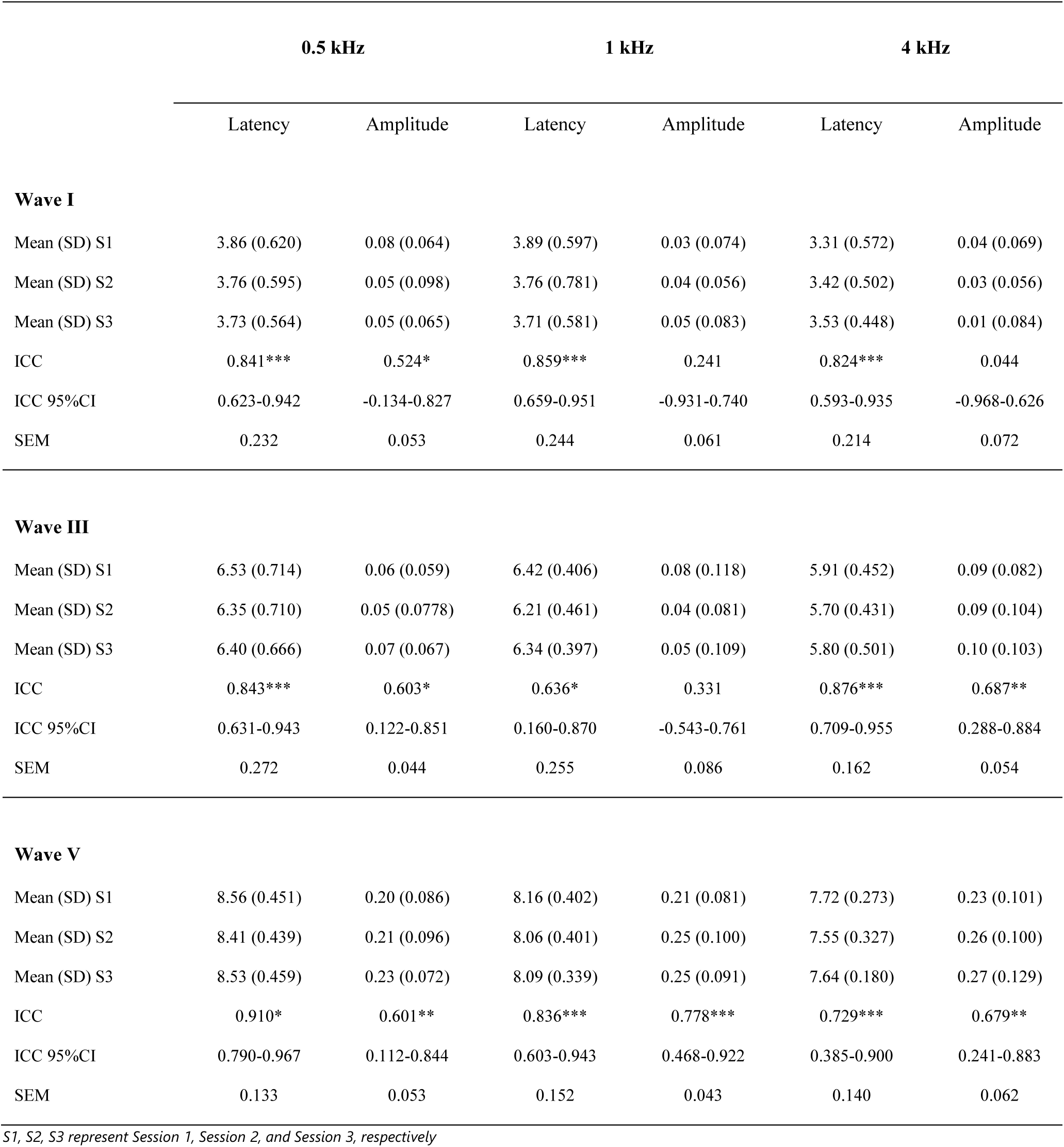
Summary of TB-ABR amplitudes (μV) and latencies (ms) per session and frequency (Hz), alongside Intraclass Correlation Coefficients (ICCs) and corresponding 95% Confidence Intervals (CIs), and Standard Errors of Measurements (SEMs). P-values for between-subjects variability are reflected as *(0.05 > p > 0.01), **(0.01 > p > 0.001), and ***(p < 0.001).

|  | 0.5 kHz |  | 1 kHz |  | 4 kHz |  |
| --- | --- | --- | --- | --- | --- | --- |
|  | Latency | Amplitude | Latency | Amplitude | Latency | Amplitude |
| **Wave I** |  |  |  |  |  |  |
| Mean (SD) S1 | 3.86 (0.620) | 0.08 (0.064) | 3.89 (0.597) | 0.03 (0.074) | 3.31 (0.572) | 0.04 (0.069) |
| Mean (SD) S2 | 3.76 (0.595) | 0.05 (0.098) | 3.76 (0.781) | 0.04 (0.056) | 3.42 (0.502) | 0.03 (0.056) |
| Mean (SD) S3 | 3.73 (0.564) | 0.05 (0.065) | 3.71 (0.581) | 0.05 (0.083) | 3.53 (0.448) | 0.01 (0.084) |
| ICC | 0.841*** | 0.524* | 0.859*** | 0.241 | 0.824*** | 0.044 |
| ICC 95%CI | 0.623-0.942 | -0.134-0.827 | 0.659-0.951 | -0.931-0.740 | 0.593-0.935 | -0.968-0.626 |
| SEM | 0.232 | 0.053 | 0.244 | 0.061 | 0.214 | 0.072 |
| **Wave III** |  |  |  |  |  |  |
| Mean (SD) S1 | 6.53 (0.714) | 0.06 (0.059) | 6.42 (0.406) | 0.08 (0.118) | 5.91 (0.452) | 0.09 (0.082) |
| Mean (SD) S2 | 6.35 (0.710) | 0.05 (0.0778) | 6.21 (0.461) | 0.04 (0.081) | 5.70 (0.431) | 0.09 (0.104) |
| Mean (SD) S3 | 6.40 (0.666) | 0.07 (0.067) | 6.34 (0.397) | 0.05 (0.109) | 5.80 (0.501) | 0.10 (0.103) |
| ICC | 0.843*** | 0.603* | 0.636* | 0.331 | 0.876*** | 0.687** |
| ICC 95%CI | 0.631-0.943 | 0.122-0.851 | 0.160-0.870 | -0.543-0.761 | 0.709-0.955 | 0.288-0.884 |
| SEM | 0.272 | 0.044 | 0.255 | 0.086 | 0.162 | 0.054 |
| **Wave V** |  |  |  |  |  |  |
| Mean (SD) S1 | 8.56 (0.451) | 0.20 (0.086) | 8.16 (0.402) | 0.21 (0.081) | 7.72 (0.273) | 0.23 (0.101) |
| Mean (SD) S2 | 8.41 (0.439) | 0.21 (0.096) | 8.06 (0.401) | 0.25 (0.100) | 7.55 (0.327) | 0.26 (0.100) |
| Mean (SD) S3 | 8.53 (0.459) | 0.23 (0.072) | 8.09 (0.339) | 0.25 (0.091) | 7.64 (0.180) | 0.27 (0.129) |
| ICC | 0.910* | 0.601** | 0.836*** | 0.778*** | 0.729*** | 0.679** |
| ICC 95%CI | 0.790-0.967 | 0.112-0.844 | 0.603-0.943 | 0.468-0.922 | 0.385-0.900 | 0.241-0.883 |
| SEM | 0.133 | 0.053 | 0.152 | 0.043 | 0.140 | 0.062 |

*S1, S2, S3 represent Session 1, Session 2, and Session 3, respectively*

TB-amplitudes generally exhibited slightly lower ICCs and higher SEMs compared to clicks. Similar to click-stimuli findings, wave I amplitudes exhibited moderate-to-very poor ICCs. For latencies, moderate-to-good ICCs with highly significant between-subject variances (*p* < .001) and small SEMs were observed for waves I- and III- and V-latencies. Figures 2 D, E, F and G display KDE-plots of wave I and V ABR-amplitudes, and -latencies, representing the distribution of individual test-retest 95%CIs for different measures.

### ENVELOPE FOLLOWING RESPONSES (EFR)

Figure 4 depicts the EFR-strength distribution for SAM- and RAM-stimuli across the three consecutive sessions.

**Figure 4:**
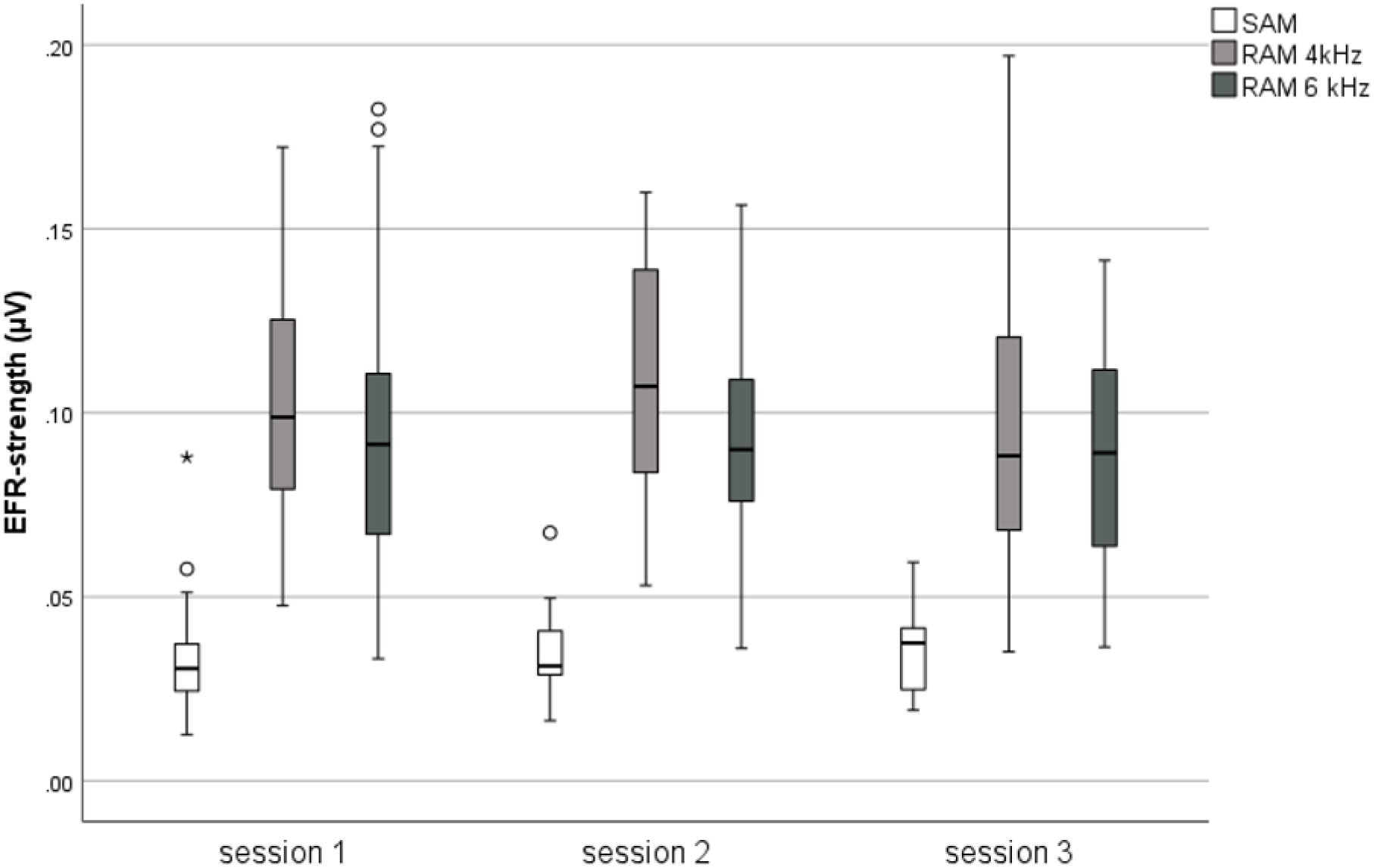
Boxplots presenting the distribution of EFR-strengths across sessions, with white, light grey, and dark grey denoting sinusoidally-amplitude modulated (SAM)-, 4 kHz Rectangularly-amplitude modulated (RAM)-, and 6 kHz RAM-EFR strengths, respectively.

Consistent with prior findings (Vasilkov et al., 2021), EFRs were stronger for the RAM stimuli than the SAM stimulus. An outlier identified in the SAM-evoked response during session two was excluded due to potential data corruption caused by a 50 Hz noise from nearby electrical equipment. No significant changes in EFR-strength were found between measurements for the SAM-stimulus [F(2, 26) = 0.066, *p* > 0.05], and RAM-stimuli; i.e. RAM 4 kHz [F(2, 28) = 0.383, *p* > 0.05] and RAM 6 kHz [F(2, 28) = 1.299, *p* > 0.05]. Moreover, the ICC demonstrated excellent test-retest reliability for both the SAM and RAM stimuli at 4 kHz and 6 kHz, yielding average measures of 0.882 (95% BI [0.708;0.959]; F(13, 26) = 7.975, *p* < 0.001), 0.950 (95% BI [0.883;0.982]; F(14, 28) = 19.355, *p* < 0.001) and 0.930 (95% BI [0.837;0.974]; F(14, 28) =14.553, *p* < 0.001), respectively. Table 7 provides detailed information on ICCs and SEMs.

**Table 7.** Summary of EFR-strengths (μV) per session and stimulus-parameter along with Intraclass Correlation Coefficients (ICCs), and corresponding 95% Confidence Intervals (CIs), and Standard Errors of Measurements (SEMs). P-values for between-subjects variability are reflected as *(0.05 > p > 0.01), **(0.01 > p > 0.001), and ***(p < 0.001).

|  | SAM | RAM 4 kHz | RAM 6 kHz |
| --- | --- | --- | --- |
| **Mean (SD) S1** | 0.03 (0.019) | 0.10 (0.038) | 0.10 (0.047) |
| **Mean (SD) S2** | 0.03 (0.015) | 0.10 (0.035) | 0.09 (0.035) |
| **Mean (SD) S3** | 0.04 (0.012) | 0.010 (0.045) | 0.09 (0.032) |
| **ICC** | 0.882*** | 0.950*** | 0.930*** |
| **ICC 95%CI** | 0.644-0.946 | 0.883-0.982 | 0.837-0.974 |
| **SEM** | 0.005 | 0.009 | 0.010 |

*S1, S2, S3 represent Session 1, Session 2, and Session 3, respectively*
*S1, S2, S3 represent Session 1, Session 2, and Session 3, respectively.*

To provide a more comprehensive perspective on observed variations in ABR- and EFR-magnitudes, both considered as potential EEG-markers of CS, Figure 5 displays individual strengths and distribution boxplots.

**Figure 5:**
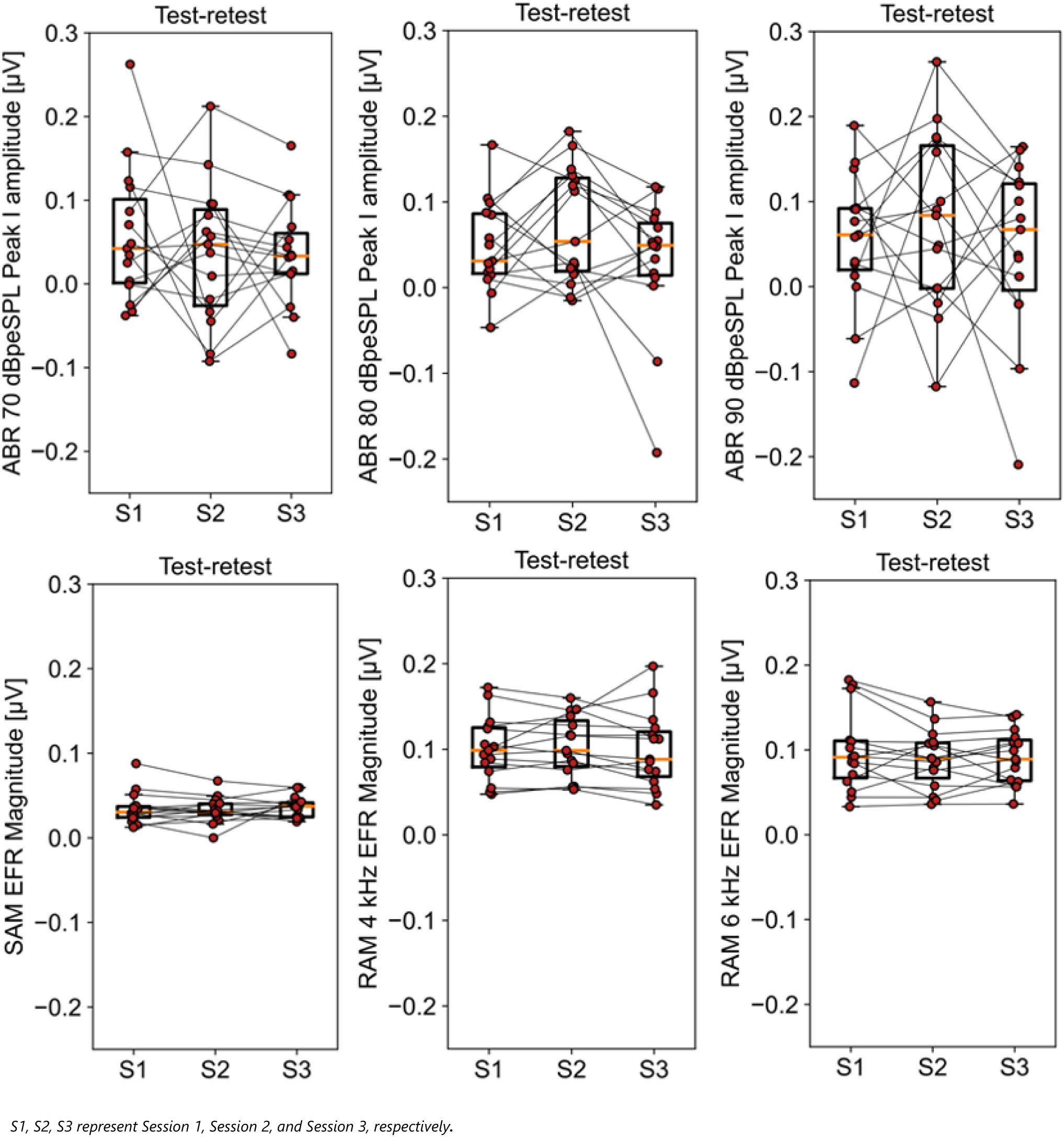
Individual ABR-amplitudes and distribution plots for click-ABRs at 70 dBpeSPL, 80 dBpeSPL, and 90 dBpeSPL (row 1), along with individual EFR-magnitudes and distribution boxplots for EFR SAM, RAM 4 kHz, and RAM 6 kHz (row 2). Horizontal lines within the boxplots denote the median ABR-amplitudes and EFR-magnitudes.

Additionally, Figure 2H presents KDE-plots for both parameters, showing the distribution of individual 95% CIs computed across three sessions. Both figures highlight the superior reliability of EFR-magnitudes over ABR-amplitudes. 95%CIs of the ICCs further confirm the higher reliability of RAM-EFRs compared to ABR-amplitudes, as their CIs demonstrate non-overlapping ranges.

## DISCUSSION

### VALIDATING THE HIGH RELIABILITY OF PURE-TONE AUDIOMETRY WITH CONSIDERATION FOR FREQUENCY-SPECIFIC VARIATIONS

Prior research has recommended using a frequency range up to 14 kHz for monitoring purposes (Rodríguez Valiente et al., 2014), as auditory thresholds beyond this frequency show substantial intra-subject threshold variability (Frank, 2001; Schmuziger et al., 2007). While the current study validated good-to-excellent ICCs for both conventional and extended high frequencies, frequency-specific disparities should be taken into consideration within clinical practice since thresholds at 6 kHz, 8kHz, and 20 kHz retained moderate-to-poor ICCs and considerably larger corresponding 95%CIs. The higher variability at 6 and 8 kHz aligns with the findings of Schlauch and Carney (2011), and may be linked to suboptimal earphone positioning or calibration methods. The increased variability at 20 kHz is likely due to standing waves, suggesting that extending the frequency range up to 20 kHz is not advisable for monitoring purposes.

The good test-retest reliability of (high-frequency) audiometry is corroborated by studies conducted by Swanepoel et al. (2010) and Ishak et al. (2011), as well as by several other investigations that employed diverse transducer models (Fausti et al., 1998; Frank, 1990, 2001; Frank & Dreisbach, 1991; Schmuziger et al., 2004).

The significant difference in 250 Hz thresholds revealed by the repeated measures ANOVA, prompts contemplation regarding the potential influence of mask-wearing on response accuracy. This is especially pertinent in lower frequency assessments, where participants reported conspicuous interference stemming from the act of breathing while wearing face masks, notably affecting their perception of very low bass tones. The observed enhancement in results during session three suggests a potential adaptation among subjects to the masking effects induced by mask-wearing.

### NEED FOR A RELIABLE SPiQ-AND SPiN TEST

SPiQ- and SPiN-tests showed no significant differences between sessions for the BB-lists, while significant threshold changes were found for the HP-lists in quiet and in noise. This learning effect was previously documented by Luts et al. (2014), highlighting a large threshold decrease occurring between the first and the second measurement which decreased to a value below 1 dB after the second list. Similar trends were observed for all language-specific tests covered in the review paper by Kollmeier et al. (2015), suggesting that the training effect might be associated more with the nature of the task and test structure rather than language-specific characteristics. However, in the current study, a training effect was observed despite the provision of two training lists. Firstly, the inclusion of two HP filtered training lists might have counteracted learning, as only BB training lists were intended. However, presenting multiple training lists extends the test duration, affecting subjects’ attention span and potentially influencing outcomes. Nevertheless, Vande Maele et al. (2021) reported significant SNR-improvements in all tested conditions, including BB, even with two BB-training lists provided. Secondly, a closed-set test format might contribute to the learning effect, as subjects could more easily learn words when both heard and visualized. The intention of displaying the possible words was to mitigate potential performance improvements across sessions, as subjects are aware of the words they may encounter from the outset. Nonetheless, the review paper of Kollmeier et al. (2015) reported a training effect in both open- and closed-set test formats for each language examined. Thirdly, within this study, participants were directed to respond in a forced-choice format to mitigate the learning effect. This was prompted by the observation that during the initial session, subjects frequently signaled non-detection of the word more swiftly, yet exhibited increasing confidence in subsequent measurements. This behavior might lead to speculation and potentially improved performance in subsequent sessions, which might indirectly contribute to better results. In sum, although further investigation is warranted, given the small study population and the potential for type I errors in the current study, caution is advised in using the matrix test in repeated measures or monitoring, due to the potential influence of learning effects.

### DPOAES EXHIBIT LARGE VARIABILITY

#### DP-GRAMS

One-way-repeated measures ANOVA indicated no significant changes in DPOAE response amplitudes between the measurements for all tested criteria and frequencies. These findings algin with a study conducted in 2010, reporting no significant differences within time-intervals up to 60 minutes (Keppler et al., 2010). However, when the time-interval extended to 7 days, a significant difference was noted, suggesting decreased reliability of DP-grams with increased time intervals (Keppler et al., 2010). Engdahl et al. (1994) and Wagner et al. (2008) noted that prolonged time intervals lead to increased standard deviations due to greater variation in middle ear pressure, room- and biological noise. Probe refitting at each session on different days further contributes to variability, as indicated by research highlighting the impact of probe replacement on the level of background noise and acoustic leakage (Beattie & Bleech, 2000; Beattie et al., 2003; Franklin et al., 1992; Keppler et al., 2010; Mills et al., 2007; Wagner et al., 2008; Zhao & Stephens, 1999).

The SEM values in this study generally surpassed those reported in other studies (Beattie et al., 2003; Franklin et al., 1992; Keppler et al., 2010; Ng & Mcpherson, 2005; Wagner et al., 2008). Several factors could account for this increased variability. Firstly, it is important to acknowledge that the testing was conducted under less-than-optimal conditions, which likely contributed to the observed discrepancies. Variability may have been influenced by probe positioning during each session, the placement of the earmuffs, and the fact that testing occurred outside a sound-attenuating booth. Additionally, subject-generated noise has the potential to impact DPOAE response amplitudes, introducing variability due to differences in patient cooperation and ear canal acoustics (Keppler et al., 2010; Wagner et al., 2008). Furthermore, equipment-related noise (Keppler et al., 2010) and recording parameters could significantly influence response amplitudes. Franklin et al. (1992) demonstrated lower reliability for DPOAE amplitudes elicited using lower primary tone level combinations (L1/L2 = 65/55 dB SPL), as used in this study, compared to higher primary level combinations (L1/L2 = 75/70 dB SPL). Another investigation, assessing two stimulus protocols, consistently found larger absolute response amplitudes for higher primary intensities, with greater variability for lower primary level combinations relative to higher intensities (Hall, 2000). Lastly, analysis strategies vary significantly across studies. In contrast to previous research, this study did not exclude responses that did not meet inclusion criteria. Instead, amplitudes were adjusted to the minimum level (i.e. the noise floor level), resulting in larger standard deviations, and consequently, larger SEMs. Additionally, the reliability varies with different inclusion criteria, as discussed earlier, and across frequencies, as depicted in Table III. The notably higher standard deviations observed at frequencies 1.0 and 1.5 kHz, compared to others, are likely attributed to low-frequency noise contaminations (Beattie et al., 2003; Keppler et al., 2010; Wagner et al., 2008; Zhao & Stephens, 1999). The increased variability in DPAOE response amplitudes at higher frequencies is probably caused by ear-canal acoustics, particularly standing waves, amplifying intrinsic variability due to differences in sound pressure at the tympanic membrane and probe microphone (Keppler et al., 2010; Mills et al., 2007).

#### DP-THRESHOLDS

Prior research has indicated the potential of estimated DPOAE thresholds to predict pure-tone thresholds (Boege & Janssen, 2002; Goldman et al., 2006; Gorga et al., 2003). However, there is a scarcity of studies investigating test-retest reliability. In the present study, a one-way repeated measures ANOVA indicated no significant alterations in DP-thresholds across measurements for all examined criteria and frequencies. Despite this, ICCs and SEMs produced notably poor outcomes, indicating a low level of reliability in this study. The highly variable nature of the I/O function among subjects and even for different stimuli at different frequencies, as highlighted by Kimberley and Nelson (1989), Hall (2000), and Harris (1990), questions the clinical utility of this method. Additionally, Harris (1990) emphasized the need for strict minimum noise requirements for reliable responses, recommending measures such as conducting DPOAE measurements in a sound-attenuating booth, setting test protocol stopping criteria for a very low noise level, and employing continuous signal averaging until the minimum noise level is reached. Popelka et al. (1993) reported that achieving a noise floor of -40 dB for recording a single I/O function may take up to 45 minutes of testing.

In conclusion, our study evaluating DPOAEs with shielded earmuffs outside a sound-attenuating booth revealed that the test-retest reliability of DPOAEs is significantly compromised under less-than-ideal measurement conditions.

### EFR MEASURES YIELD BETTER TEST-RETEST RELIABILITY, RELATIVE TO THE ABR

The present study showed no significant changes for waves I, III and V between the three different test sessions. These results align with a 2018 study, showing no significant changes in click- and speech-evoked brainstem responses across test sessions (Bidelman et al., 2018). Additionally, Munjal et al. (2016), evaluating the reliability of the absolute latency of waves I, III and V and interpeak latencies, showed good test-retest reliability for all response parameters, except for the absolute latency of wave I.

The analyses of ICCs and SEMs unveiled several trends among the different waves and stimuli. Firstly, a higher within-subject reliability was found for wave V relative to wave I. These results are in line with the study of Lauter and Karzon (1990), who reported low level of consistency across subjects for wave I of ABR. Sininger and Cone-Wesson (2002) have shown that peripheral hearing and testing parameters, amongst others ambient noise and minimal wax in the external auditory canal, can affect the latency of wave I in ABR-measurements. Secondly, wave I may potentially be reduced due to CS and OHC-damage, while wave V may be enhanced due to central gain mechanisms (Auerbach et al., 2014; Gu et al., 2012; Schaette & McAlpine, 2011). Therefore, the interpretation of wave I-amplitudes and latencies requires some caution.

In addition to greater reliability for wave V relative to wave I, the present study also retained smaller ICCs and larger SEMs for click-amplitudes compared to click-latencies, consistent with Bidelman et al. (2018). Negligibly small intra-subject variability in ABR latencies are in addition in agreement with previous studies of Edwards et al. (1982) and Oyler et al. (1991). Firstly, evoked potential amplitudes are susceptible to nonbiological factors, such as electrode impedance and orientation relative to source generators. This suggest that the amplitude might be a poor metric for reliably assessing subtle changes in ABR-measurements with certain experimental manipulations, including noise exposure, ototoxicity, age, and training (Bidelman et al., 2018). The use of ear canal tiptrodes, as opposed to scalp mounted electrodes could result in higher reliability since the recording site has moved closer to the generator of wave I, specifically the auditory nerve. This assumption aligns with the study of Bauch and Olsen (1990), showing increased wave I-amplitudes with ear canal tiptrodes compared to mastoid electrodes, and Bieber et al. (2020), reporting good-to-excellent wave I and wave V amplitude ICCs when measured from the ear canal. However, the study of Prendergast et al. (2018) demonstrated only a small increase in reliability for waves I and V when using canal tiptrodes compared to mastoid electrodes. The benefits for the summation potential however, were greater. In sum, while wave I has proved valuable, particularly in research studies, as a more direct measure of peripheral auditory function, our study revealed low amplitude ICCs, casting doubt on the feasibility of clinical waveform interpretation under the specified conditions. Prior investigations with good-to-excellent test-retest reliability often extended their test durations, incorporating up to 10000 sweeps and/or or automated peak- and trough-picking procedures (Bieber et al., 2020; Guest et al., 2019; Prendergast et al., 2018), suggesting considerable advantages of automated peak-picking algorithms in clinical practice. Moreover, stimulus levels were frequently elevated, reaching 115.5 dB peSPL in the study by Prendergast et al. (2018). Finally, it is essential to acknowledge that the relatively modest size of our study population and the testing environment within hospital settings could have influenced the observed results.

When comparing broadband clicks to toneburst stimuli in our study, it is evident that clicks evoke larger responses and serve as slightly more reliable biomarkers. This is likely because broadband clicks activate more auditory nerve fibers simultaneously, potentially resulting in larger and more robust responses. Subsequently, TBs are identified as less clearly detectable peaks, indicating higher interrater variability and therefore lower overall reliability. This hypothesis is supported by the generally smaller amplitudes and longer latencies in TB-responses, which likely stem from the narrower basilar-membrane stimulation (Gorga et al., 1988; Rasetshwane et al., 2013). Additionally, the impact of prolonged latencies and reduced amplitudes with narrower BM stimulation is more pronounced at lower frequencies compared to higher frequencies (500 Hz vs 4 kHz), primarily due to cochlear wave dispersion (Rasetshwane et al., 2013).

As noise-induced CS primarily targets AN fibers with high thresholds, and phase locking to temporal envelopes is in addition particularly strong in these fibers, the EFR-strength could potentially be a more robust measure, relative to ABR-amplitudes (Vasilkov et al., 2021).

Additionally, phase information can be extracted from EFRs, and measures of phase-locking values might be less susceptible to anatomical variations in humans (Gorga et al., 1988), generally interfering amplitude measures. These findings align with Bidelman et al. (2018), who reported that Frequency Following Responses generally showed higher test-retest reliability compared to conventional click-evoked ABRs. Furthermore, our study’s results, which indicated favorable ICCs and non-overlapping corresponding 95%CIs between ABR-amplitudes and RAM-EFR strengths, align with this observed trend. However, Guest et al. (2019) reported only minor differences in reliability between ABR amplitudes and EFR strengths. Although their experimental setup resembled ours in terms of the number of presentations (ranging from 5200-56000 versus 4000) and click levels (90, 96, and 102 dB peSPL versus 70, 80 and 90 dBpeSPL), they employed automated peak-picking algorithms to select peaks, which likely improved to reliability of the ABR results.

## CONCLUSION

In the pursuit of identifying noninvasive early markers of noise-induced SNHL in humans, various measures have been explored. However, comprehensive studies evaluating test-retest reliability of multiple measures and stimuli within a single study remain limited, and a standardized clinical protocol encompassing robust noninvasive early markers of SNHL has not yet been established. Addressing these gaps, this exploratory study aimed to explore the intra-subject variability of various potential noninvasive EEG-biomarkers of CS and other early indicators of SNHL within the same individuals. While pure-tone audiometry has confirmed good reliability in this study, caution is advised when extending the frequency range beyond 16 kHz. The observed learning effect in the speech-sentence test emphasizes the need for caution when employing the matrix sentence test in repeated measurements. The variability noted in DPOAEs highlights the importance of consistent ear probe replacement, meticulous measurement techniques, and optimal testing conditions to minimize variability in DP-amplitudes, indicating that DPOAE test-retest reliability is significantly compromised under less-than-ideal conditions. Regarding auditory evoked potentials, the study found that EFRs exhibited greater reliability compared to ABRs when manually selecting the ABR-waveforms.

## Acknowledgements

The authors would like to thank Eef De Wilde and Ellen Sabau for their significant contribution in the data-collection.

## Data Availability Statement

The datasets generated during and/or analyzed during the current study are not publicly available due to ethical restriction but are available from the corresponding author on reasonable request.

## Ethics approval Statement

This study received approval from the UZ Gent ethical committee (BC-05214) and adhered to the ethical principles outlined in the Declaration of Helsinki. All participants were informed about the testing procedures and provided an informed consent.

